# Methods for analytical validation of novel digital clinical measures: A simulation study

**DOI:** 10.1101/2024.11.29.24318211

**Authors:** Simon Turner, Chen Chen, Rolando Acosta, Eric J Daza, Lysbeth Floden, Joss Langford, Leif Simmatis, Berend Terluin, Piper Fromy

## Abstract

Analytical validation (AV) is a crucial step in the evaluation of algorithms that process data from sensor-based digital health technologies (sDHTs). AV of novel digital measures can be complicated when reference measures with direct correspondence are not available. To address this, we conducted a simulation study. Data was simulated assuming a latent physical ability trait, indirectly accessed through an sDHT-derived target measure collecting step count data, and the items of a clinical outcome assessment measuring self-reported physical activity. We quantified the ability of two methods to assess the latent relationship between reference and target measures: the Pearson Correlation Coefficient (PCC) (a classical, simple method frequently used for AV), and factor correlations from a two-factor confirmatory factor analysis (CFA) model (a more novel approach to AV, able to capture more complex relationships). Additionally, three multiple linear regression models were used to evaluate if multiple clinical outcome assessment reference measures can more completely represent a target measure of interest.

Our findings show that PCC was more stable, easier to compute, and relatively robust with respect to violations of parametric assumptions than CFA, particularly with small sample sizes. However, CFA was less biased than PCC in all scenarios investigated. We demonstrate that using both PCC and CFA allows a researcher to be more confident that their AV results reflect the true relationship between the sDHT-derived target measure and a reference measure. Finally, regression results suggest that incorporating multiple reference measures with more frequent collection time points can provide a more complete presentation of the sDHT’s analytical validity.

Novel digital measures are being developed at an accelerating pace and promise to revolutionize public health, patient care, and medical product development. Our findings provide investigators with crucial information for choosing appropriate methods to perform rigorous AV of these novel measures.

## Introduction

For a sensor-based digital health technology (sDHT) to support scientific and clinical decision-making, its sensor(s) must be evaluated by verification, any algorithm(s) must be analytically validated, and clinical validation must be conducted to ensure it measures a relevant functional state in a specific context of use, as codified in the V3+ framework by the Digital Medicine Society (DiMe) [1,2]. Analytical validation (AV) ensures an sDHT algorithm measures, detects, or predicts physiological or behavioral metrics by comparing its output to a rigorous reference standard.

Prior work by Bakker et al. [3] developed a framework for selecting a fit-for-purpose reference measure when conducting AV. In the case of novel digital measures, defined as those measuring a construct that was previously inaccessible or a known construct in a previously infeasible environment, a fit-for-purpose reference measure may not exist. In those cases, often the only available options are clinician-reported outcomes (ClinROs) and patient-reported outcomes (PROs). These measures are popular as surrogate endpoints in clinical research [4,5], especially in therapeutic areas such as neurodegenerative disease [6], mental health, and cardiology [7,8], where meaningful and sensitive measures are challenging to define or collect [9,10]. However, ClinROs and PROs are collected subjectively and episodically, and may be only moderately related to a measure derived from the intensive longitudinal data [11] commonly collected by a sDHT.

AV with such reference measures is complicated. Their intrinsic measurement error makes comparison to sDHT-derived target measures challenging, and they typically do not measure the desired concept or construct directly. Assessment of direct correspondence (i.e., when the reference measure collects the same data as the target measure, in the same units, with the same resolution and at the same point in time) between the target and reference measures is often infeasible. That said, total replication of an existing reference measure’s output is often undesirable. Rather, one may seek to improve upon such reference measures by instead using a sDHT to measure the desired construct.

As an example of this complicated situation, consider methods designed to assess analytical validity of sDHT-derived measures in Parkinson’s disease. Comparison against reference measures such as the Movement Disorder Society-Unified Parkinson’s Disease Rating Scale (MDS-UPDRS) [12] is often needed. MDS-UPDRS is a battery of assessments that combine PROs and ClinROs. It is widely used in both research and care of Parkinson’s disease, but is known to be subjective, collected infrequently, and suffers from high within-subject variability and low test-retest reliability [13]. Results of comparisons between sDHT measures and MDS-UPDRS are often moderate to poor, with potentially substantial individual variability in sDHT performance relative to the reference measure. However, such results do not definitively prove or disprove the validity of sDHT measures. It is hard to know what is a good performance value and it is difficult to pinpoint the cause of a lack of good performance, as it could stem from a lack of agreement at the latent level, measurement errors, or a lack of direct correspondence between the measures. Moreover, the community has shifted away from optimizing sDHT development to replicate MDS-UPDRS because it would only address the issue of infrequent data collection, while still leaving researchers with a noisy, potentially unreliable measure. Therefore, the development of sDHT measures focuses on objectively assessing similar concepts as the MDS-UPDRS, even if they may not correlate well with MDS-UPDRS.

Given these challenges, selection of appropriate reference measures as defined by Bakker et al. [3], and selection of appropriate AV methodology are crucial. Specifically, methods for characterizing convergent validity – how well responses across conceptually similar tests, instruments, or scales relate to each other, can be employed. These focus on assessing the relationship between the sDHT algorithm outputs and reference measures.

Convergent validity methods assume that a latent trait of a given construct exists. A latent trait represents a true underlying status that the construct tries to quantify, such as severity of a disease or physical ability to conduct daily activities. Crucially, although a latent trait is unseen and immeasurable, it influences a given person’s behavior. This means that an individual’s latent trait can be estimated using *indicators* of that trait.

Borrowing concepts from psychometric research, from which the theories of convergent validity emerged, the indicators are typically questionnaire items related to the topic under investigation [14]. With an sDHT-derived measure, the indicator of the underlying construct is the output of the algorithm(s). Statistical methods such as correlation, concordance analysis, and regression analysis are commonly employed in this scenario. If we consider the repeated measures of a sDHT to be different indicators of a latent trait, then confirmatory factor analysis (CFA) offers an appealing alternative, because of its promise to extract and obtain the correlation between latent traits, instead of correlation between observed measures.

The properties of these methods are well understood in the statistical community. However, their behaviors in the context of characterizing the relationship between a sDHT-derived measure and a reference measure have not been explored extensively to the best of our knowledge. This work focuses on exploring these properties in a simulation study.

Simulation is a foundational scientific tool to evaluate properties of statistical methods because the “truth” is known. More pertinently, it is useful in the context of digital measure development. Data in the medical research field are scarce, and it is extremely challenging to gather enough data to experiment with all the scenarios of interest. Simulation provides easy access to a large data pool, and enables researchers to control and vary parameters that influence the manifestation of observed values from latent traits.

Our paper has two aims. Firstly, we assess and compare the statistical properties of two methods that are used to evaluate convergent validity: Pearson correlation coefficients of observed measures, and the model-implied correlation coefficient of the factors derived by a two-factor CFA model. Secondly, we seek to understand if or how multiple reference measures can provide more information for characterizing a target measure’s convergent validity.

## Methods

We evaluated the performance of statistical methods when validating a digital measure against one or a combination of reference measures representing COAs. The simulation assumes count data from the sDHT, representing a record of events occurring, at the summary level. By this, we mean that we simulate the total count of events occurring on each day, rather than simulating the total number of events taking place in epochs during the day and then summing these values. The canonical example motivating our choices in this simulation is that of physical activity, where the sDHT data represents the count of steps taken during a particular day. The latent trait is assumed to be an underlying measure of physical ability that fluctuates daily during the seven days of the study. The COAs are assumed to be measures of patient- or clinician-reported physical activity. The data generation mechanism (DGM) assumed an underlying latent trait being indirectly accessed through both the sDHT algorithm and the items of each COA.

### Data generation mechanism (DGM)

We simulated seven days of data from the sDHT’s algorithm (i.e., the target measure), and data from multiple COAs (i.e., the reference measures). Figure 1 depicts a summary of the DGM we used.

**Figure 1.**
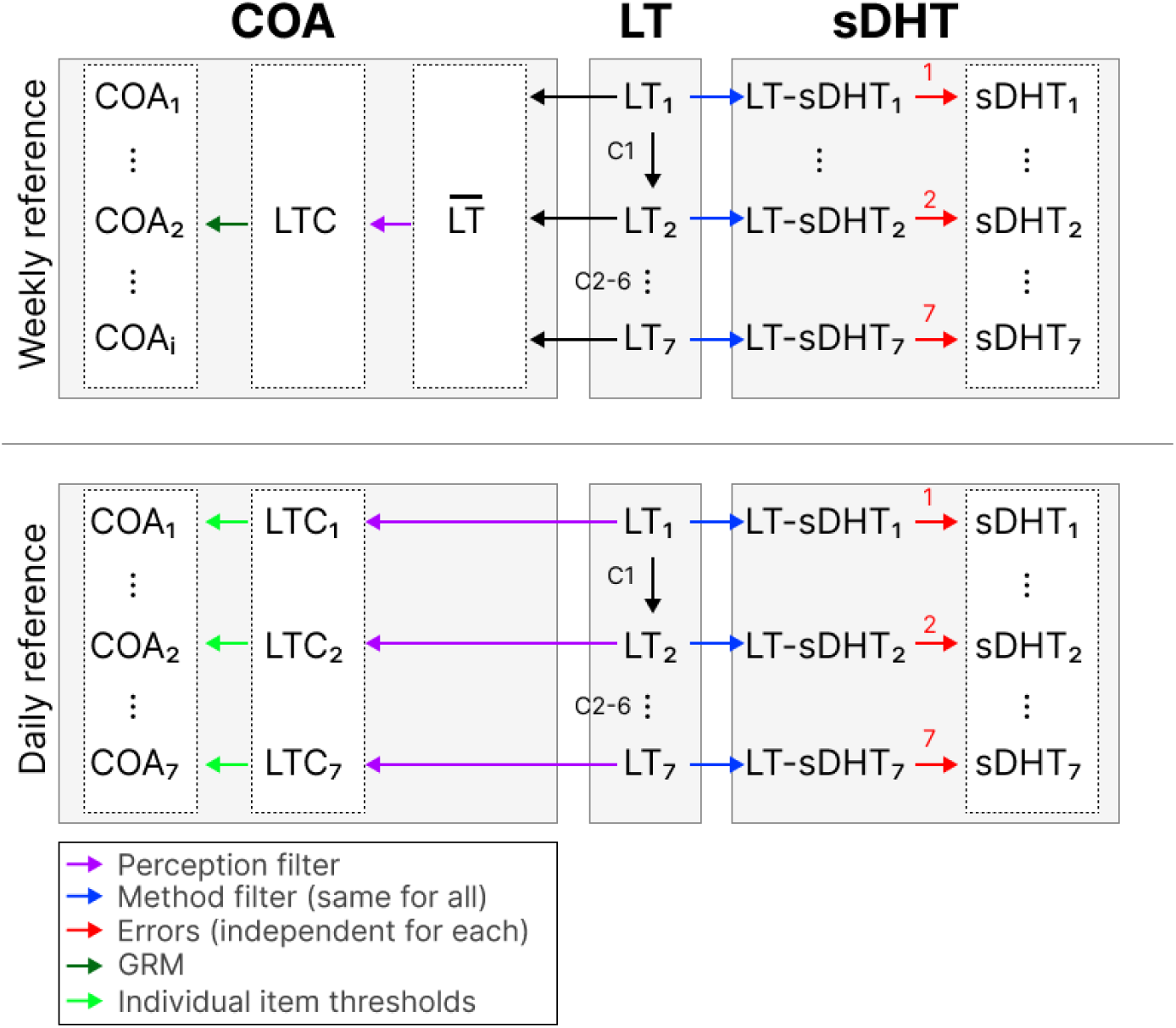
A diagram depicting the DGM used in this study, illustrating the relationship between the latent and observable variables for the target measure (sDHT-based) and the different reference measures (COA-based). C{1-6} = fluctuation factors 1-6; COA = clinical outcome assessment; GRM = graded response model; LT = latent trait; LTC = latent trait for the COA; LT-sDHT = latent trait for the sDHT.

**Table 1.** Notation used in the DGM.

| Notation | Description |
| --- | --- |
| $\theta_{ij}$ | The value of the latent variable for individual j on day i of the study |
| $\theta'_{ij}$ | The value of the “method-filtered” latent variable for individual j on day i of the study |
| $\lambda_{ij}$ | The mean of the Poisson distribution used to generate individual j’s target measure data from day i |
| $\lambda_0$ | The base rate for the target measure (i.e., , the mean of the target measure for a latent variable value of zero) |
| $\lambda_{\text{effect}}$ | The effect of the latent variable on the daily mean of the target measure |
| $\epsilon_{ij}$ | The measurement error of the sDHT |
| $\sigma_{ij}^2$ | The magnitude of the sDHT measurement error (MEM) |
| $\theta_j$ | The mean value of the latent trait over the seven days for individual j |
| $\overline{\theta_j^P}$ | Individual j’s “perception-filtered” latent trait for the weekly PRO |
| $DHT_{ij}$ | The observed count data for individual j on day i |
| $wPRO_j$ | Rescaled total score from the weekly PRO for individual j |
| $\theta_{ij}^P$ | Individual j’s perceived latent trait value for the daily PRO on day i |

### Simulating the latent variables

The latent variable for each individual on day one was simulated from a standard Normal distribution. The latent variable for each subsequent day was simulated by adding a fluctuation factor to the latent variable from the previous day, and then rescaling to give a sample variance of 1.0 for the data across all individuals on that day. The fluctuation factor was taken from a Normal variable with mean 0 and standard deviation (SD) 0.30.

### Simulating the target measure data

For each individual, we simulated seven counts, representing seven days of activity, to mimic reliable data across a spectrum of populations and contexts-of-use [15,16,17]. Data were to be assessed on consecutive days; effects of weekdays and weekends were not specified. \

To mimic the imperfect ability of the target measure to observe the latent trait, Gaussian noise with mean 0 and SD 0.5 was added to θ*_ij_* to obtain θ*_ij_*’ for individual] on day i. This “method filter” did not vary between days.

To simulate the count data, a random value was drawn from a Poisson distribution with mean λ*_ij_*, where λ*_ij_* was calculated as:

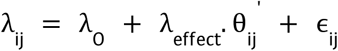

Here, λ_0_ is the hypothesized mean of the target measure for a latent variable value of zero (i.e., an individual from the population with mean physical ability). This term is independent of an individual’s latent ability. In this study, λ_0_= 10 000. The λ_effect_ term represents the proportional effect of an individual’s latent physical ability on their expected target measure count. In essence, λ_effect_ scales an individual’s “filtered” latent trait. In this study, λ_effect_= 1 250. The final term (ɛ_ij_) represents measurement error, and is Normally distributed with mean 0 and SD σ^2^_ij_. Henceforth, σ^2^_ij_ will be called the sDHT-measurement error magnitude (MEM), and will be varied as a simulation parameter.

The values of the above parameters were chosen to reflect a correlation matrix representative of real-world physical activity digital measure data [18]. Each Poisson mean was constrained <as λ > 0.

The above approach was used for both research aims investigated in this study. The observed count data for individual] on day i is henceforth denoted *DHT_ij_*.

### Simulating the reference measure data

#### Primary reference measure: Weekly PRO

The reference measure simulated for the primary aim of the study is modeled as a four-response, twelve-item PRO, administered at the end of the seven day period. Since each individual is assumed to recall information across the entire study period when responding to the PRO, the value from each day of the latent variable will impact their answers. To model this, we used 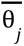, the mean of each individual’s latent traits over the seven simulated days, as the starting point.

We assumed that an individual has an imperfect ability to perceive their 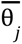 value when recalling their activities during the previous seven days. To simulate this, we added *p* random noise generated from a Normal distribution with mean 0 and SD 1.0. Let 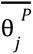 denote individual j’s “perception-filtered” latent trait for this reference measure.

An individual’s 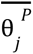 value was used to simulate their responses to a twelve-item PRO with four-response options. The item scores were simulated using an item response theory (IRT) graded response model (GRM). The difficulty threshold parameters took the following fixed values across each individual:

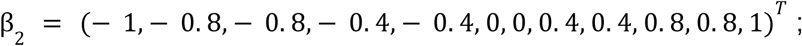

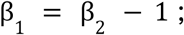

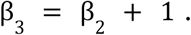

The discrimination parameter α also took a fixed value across each individual. The value of α was chosen such that the resulting simulated PRO had an approximate reliability of 0.8. A total score was then derived as the sum of the item scores, rescaled to a 0-100 scale. These total scores are the final reference measure data that were compared to the target measure data, and are denoted by *wPRO.* for individual].

#### Secondary reference measures: Weekly ClinRO and daily PRO

The secondary aim of this study explores whether introducing multiple reference measures can help to provide more information in assessing a digital measure’s convergent validity. The additional reference measures simulated were a five-point, seven-item, weekly ClinRO and a single-item, daily five-point PRO representing a patient’s global impression of severity.

The weekly ClinRO was simulated in a similar way to the weekly PRO. We simulated an individual’s imperfect recall over the seven days by applying a perception filter to 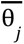, adding random noise generated from a Normal distribution with mean 0 and SD 1.0. We then used an IRT GRM to simulate each individual’s responses to the seven items on the five-point scale. The difficulty threshold parameters took the following fixed values across each individual:

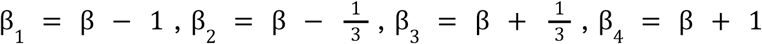

Here, β = (— 0. 75, — 0. 5, — 0. 25, 0, 0. 25, 0. 5, 0. 75)*^T^*.

The discrimination parameter α also took a fixed value across each individual, and was chosen such that the resulting simulated ClinRO had an approximate reliability of 0.7. A total score was calculated as the sum of the item scores, rescaled to a 0-100 scale. These rescaled total scores were the variables analyzed in our study.

The single-item daily PRO was simulated using the approach in Griffiths et al [19] Firstly, it was assumed that the response of individual] to a single item (on a five-point, 0-4 scale) on day i depends on θ*_ij_*.

To simulate the imperfect ability of an individual to perceive their θ_value, we added random noise generated from a Normal distribution with mean 0 and SD 0.5. This “perceived latent trait” value is denoted θ*_ij_^p^*, and is assumed to exactly determine (i.e., without noise) the response of individual] on day i.

It was assumed that on average, the thresholds at which an individual gives a particular response were -1.5, -0.5, 0.5, and 1.5. For example, a perceived latent trait value of -1.6 should on average result in a score of 0; a perceived latent trait value of 0.8 should on average result in a score of 3, and so on.

Finally, we further assume that these score threshold values vary slightly based on an individual’s own perception; for example, what one individual may consider to be “vigorous” physical activity, another individual may consider to be “moderate” physical activity. Therefore, each individual’s score thresholds were generated from Normal distributions with means -1.5, -0.5, 0.5 and 1.5, each with SD 0.075. An individual’s θ*_ij_^p^* value was then compared against their score thresholds to determine their daily PRO score on day i, and this process was repeated for each of the seven days in the study.

### Statistical methods

For the primary aim of the study, Pearson’s correlation and CFA factor correlation were used to assess the strength of the relationship between the target and the weekly PRO reference measure.

Firstly, the true relationship between the target measure and the reference measure was calculated. This was done by computing 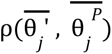 (i.e., the linear correlation between the mean of the sDHT “method-filtered” latent values and the “perception-filtered” mean of the latent values).

Two estimates of the true relationship were computed. Firstly, the Pearson correlation coefficient (PCC) between the weekly mean of the observed target measure values (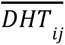) and the observed reference measure values *(wPRO_j_)* was computed. Secondly, a two-factor CFA model with correlated factors was created, and the model-implied correlation coefficient of the factors was obtained. To construct the CFA model, each item from the reference measure was loaded as an indicator onto a “reference measure factor”, and each day of the target measure data was loaded as an indicator onto a separate “sDHT” factor. The standardized value of the factor covariance was computed, and these were the factor correlation variables analyzed in our study. Figure 2 depicts the path diagram for the two-factor CFA model used in this simulation.

**Figure 2.**
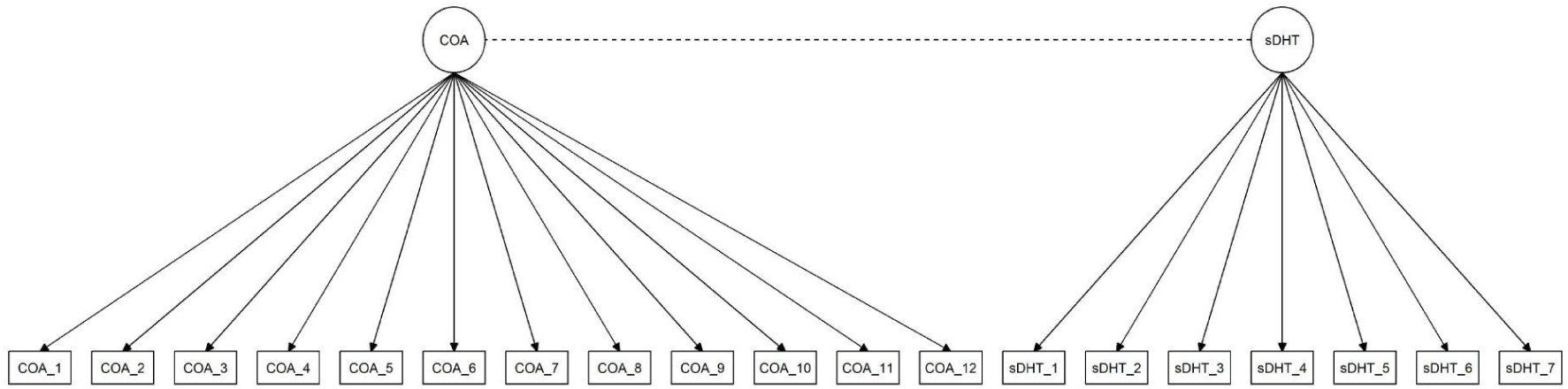
Path diagram for the CFA model used in investigating the primary aim of the study.

For the secondary aim of the study, simple linear regression (SLR) and multiple linear regression (MLR) were used to assess the impact of using more than one reference measure to conduct AV. Specifically, these methods were used to assess how much useful information can be added when using more than one reference measure.

For each regression model, 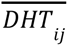 was used as the dependent variable. Ordinary least squares was used for the SLR model, with *wPRO_j_* as the predictor variable. MLR was employed in three different ways: Firstly, by introducing the weekly ClinRO as a predictor variable alongside the weekly PRO (known as the “weekly COAs model”); secondly, by introducing the mean of the daily PRO values as a third predictor variable alongside the weekly COAs (known as the “daily means” model); and lastly, by instead introducing the individual daily PRO scores as seven distinct predictor variables alongside the weekly COAs (knowns as the “daily distinct” model).

For each linear regression model used, a coefficient of determination was computed.

In the SLR case, this was the standard R^2^ statistic. In the MLR case this was the adjusted 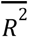, denoted R and calculated using the formula 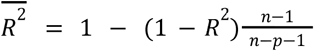, where n is the number of observations and p is the number of reference measures.

### Parameters of the simulation

The simulation varied the number of included COA measures and other parameters, in order to assess their impact on the performance of the statistical methods investigated. The following parameters were tested in a fully factorial design:

1. sample size (N = 35, 100, 200, 1000)
2. number of repeated assessments of the target measure included in the analysis (RA = 1, 3, 5, 7)
3. Missing data rate (MDR = 0, 0.10, 0.25, 0.40)
4. Missing data mechanism (MDM - No missing data, missing completely at random (MCAR), missing not at random (MNAR)) [20,21]
5. MEM 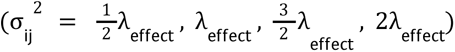

For a more detailed explanation of the missingness (i.e., missing data pattern) mechanisms employed, and of the implementation of fewer repeated assessments of the target measure, in the simulation, please see the supplementary materials 1.

### Performance measures

#### Primary aim of the study

To assess the ability of PCC and CFA factor correlation to observe the underlying relationship between the target and primary reference measure, the empirical bias of both methods was computed. The empirical bias of a method (that can be defined in terms of statistical estimators) is defined as the difference between a.) the true relationship between the target measure and the reference measure, and b.) the sample-based estimate of this true relationship from that method. The empirical bias for PCC is the difference between 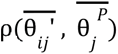 and 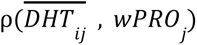. For CFA, the empirical bias is the difference between 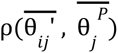 and the factor correlation estimate.

To assess the precision of the above methods, for each simulation condition the empirical standard error (empSE) of the PCC and the CFA factor correlation were computed. EmpSE is defined as the square root of the variance of each method’s correlation estimates.

In addition, for the CFA model, four model fit statistics were calculated to evaluate the goodness of fit between the proposed model and the observed data. The Comparative Fit Index (CFI), Tucker-Lewis Index (TLI), Root Mean Square Error of Approximation (RMSEA), and Standardized Root Mean Square Residual (SRMR) were the fit statistics used. The rate that the model fit statistics suggested an acceptable model fit were reported for each simulation condition. Interpretation of fit statistics were guided by the following thresholds [22,23]

- CFI and TLI acceptable fit: values ≥ 0.90.
- RMSEA and SRMR acceptable fit: values < 0.08.

The convergence, or failure rates 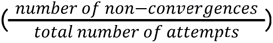, for the CFA model were reported for each simulation condition.

#### Secondary aim of the study

To assess the ability of the MLR models to recover more information about the underlying target measure-reference measure relationship than the SLR model, the change in information added, denoted *ΔR*^2^, was computed for each MLR model. *ΔR*^2^ is defined as the difference between the MLR adjusted R2 estimate and the SLR R^2^ estimate; in other words, 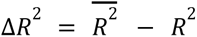.

To assess the precision of each regression method, for each simulation condition, the empSE of the SLR model and each MLR model were computed.

Finally, any failures that occurred in each of the regression models were reported.

The number of simulation repetitions (*n_sim_*) was chosen to ensure that the Monte Carlo Standard Error (MCSE) of the empSE for each statistical method was less than 0.025. Therefore *n_sim_* must satisfy the inequality 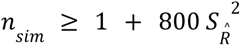, where 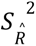 is the sample variance of the estimates [24,25]. Based on the results of a small pilot simulation, n_sim_ = 500 is sufficient to achieve the desired MCSE. The MCSEs of each model were reported for each simulation condition.

## Results

### Primary aim of the study

Table 3 depicts a summary of the mean empirical bias and mean empSE for the PCC and CFA methods under the influence of different simulation conditions, as well as an overall summary across all conditions.

**Table 3.** Mean (SD) of the empirical bias and empirical SE for each method, grouped in various ways.

|  | Mean (SD) Empirical Bias |  | Mean (SD) Empirical SE |  |
| --- | --- | --- | --- | --- |
|  | *Pearson R of raw measures* | *CFA factor correlation* | *Pearson R of raw measures* | *CFA factor correlation* |
| **Overall** | -0.160 (0.107) | 0.012 (0.111) | 0.076 (0.046) | 0.100 (0.071) |
| By Sample Size |  |  |  |  |
| 35 | -0.159 (0.145) | 0.024 (0.189) | 0.140 (0.029) | 0.193 (0.071) |
| 100 | -0.161 (0.104) | 0.014 (0.093) | 0.081 (0.017) | 0.103 (0.030) |
| 200 | -0.161 (0.089) | 0.008 (0.065) | 0.057 (0.011) | 0.072 (0.021) |
| 1000 | -0.161 (0.077) | 0.002 (0.029) | 0.025 (0.005) | 0.032 (0.009) |
| By MEM |  |  |  |  |
| $\frac{1}{2}\lambda_{\text{effect}}$ | -0.079 (0.061) | 0.013 (0.066) | 0.068 (0.040) | 0.073 (0.043) |
| $\lambda_{\text{effect}}$ | -0.132 (0.080) | 0.018 (0.08) | 0.073 (0.044) | 0.083 (0.048) |
| $\frac{3}{2}\lambda_{\text{effect}}$ | -0.190 (0.097) | 0.016 (0.112) | 0.079 (0.047) | 0.104 (0.068) |
| $2\lambda_{\text{effect}}$ | -0.241 (0.108) | 0.001 (0.161) | 0.083 (0.049) | 0.139 (0.097) |
| By number of sDHT assessments |  |  |  |  |

|  | **Mean (SD) Empirical Bias** |  | **Mean (SD) Empirical SE** |  |
| --- | --- | --- | --- | --- |
|  | *Pearson R of raw measures* | *CFA factor correlation* | *Pearson R of raw measures* | *CFA factor correlation* |
| 1 | -0.217 (0.132) | N/A | 0.093 (0.055) | N/A |
| 3 | -0.160 (0.101) | 0.010 (0.121) | 0.076 (0.044) | 0.112 (0.074) |
| 5 | -0.138 (0.086) | 0.012 (0.108) | 0.070 (0.040) | 0.097 (0.071) |
| 7 | -0.126 (0.077) | 0.014 (0.102) | 0.066 (0.037) | 0.091 (0.069) |
| By missing data mechanism |  |  |  |  |
| None | -0.145 (0.095) | 0.016 (0.081) | 0.067 (0.038) | 0.082 (0.053) |
| MCAR | -0.159 (0.111) | 0.017 (0.116) | 0.082 (0.049) | 0.106 (0.074) |
| MNAR | -0.167 (0.106) | 0.006 (0.114) | 0.073 (0.043) | 0.100 (0.074) |
| By missing data rate |  |  |  |  |
| 0 | -0.145 (0.095) | 0.016 (0.081) | 0.067 (0.038) | 0.082 (0.053) |
| 0.10 | -0.151 (0.099) | 0.016 (0.090) | 0.070 (0.041) | 0.089 (0.058) |
| 0.25 | -0.162 (0.107) | 0.013 (0.108) | 0.076 (0.045) | 0.099 (0.068) |
| 0.40 | -0.177 (0.118) | 0.004 (0.141) | 0.086 (0.052) | 0.121 (0.089) |

#### Empirical Bias

Overall, PCC was negatively biased and CFA was positively biased, but CFA was the less-biased of the two methods. The same behavior was observed when splitting the bias results by any single simulation parameter.

When considering the impact of any single simulation parameter on the mean empirical bias, the negative bias of PCC increased as the MEM and missing data rate increased. The missing data mechanism had only a minor impact on the mean empirical bias of PCC, and the sample size had essentially no impact. The negative bias of PCC decreased as the number of repeated assessments increased.

For the CFA method, any single simulation parameter had little impact on the mean empirical bias. At the maximum MEM level the CFA method was slightly less biased than for lower MEM values; the same behavior was observed for the maximum missing data rate. Data that were MNAR produced a slightly smaller magnitude of the bias when compared to no missing data or MCAR data. Varying the number of repeated target measure assessments included had no discernible impact on the mean empirical bias. When increasing the sample size, there was a very slight trend for the mean empirical bias of CFA to decrease, and this was the strongest of the trends exhibited when considering a single simulation condition.

When zooming into a given simulation condition (no missing data and 7 repeated assessments per subject; Figure 4), sample size had less of an impact on the average trend of empirical bias than MEM, while the dispersion of empirical bias appeared to decrease as sample size increased. There was a pattern of increasing underestimation from PCC as MEM increased. In contrast, CFA slightly overestimated the bias, but the average trend was not greatly affected by the varying measurement error. This pattern was repeated in the 3-assessment and 5-assessment case.

**Figure 4.**
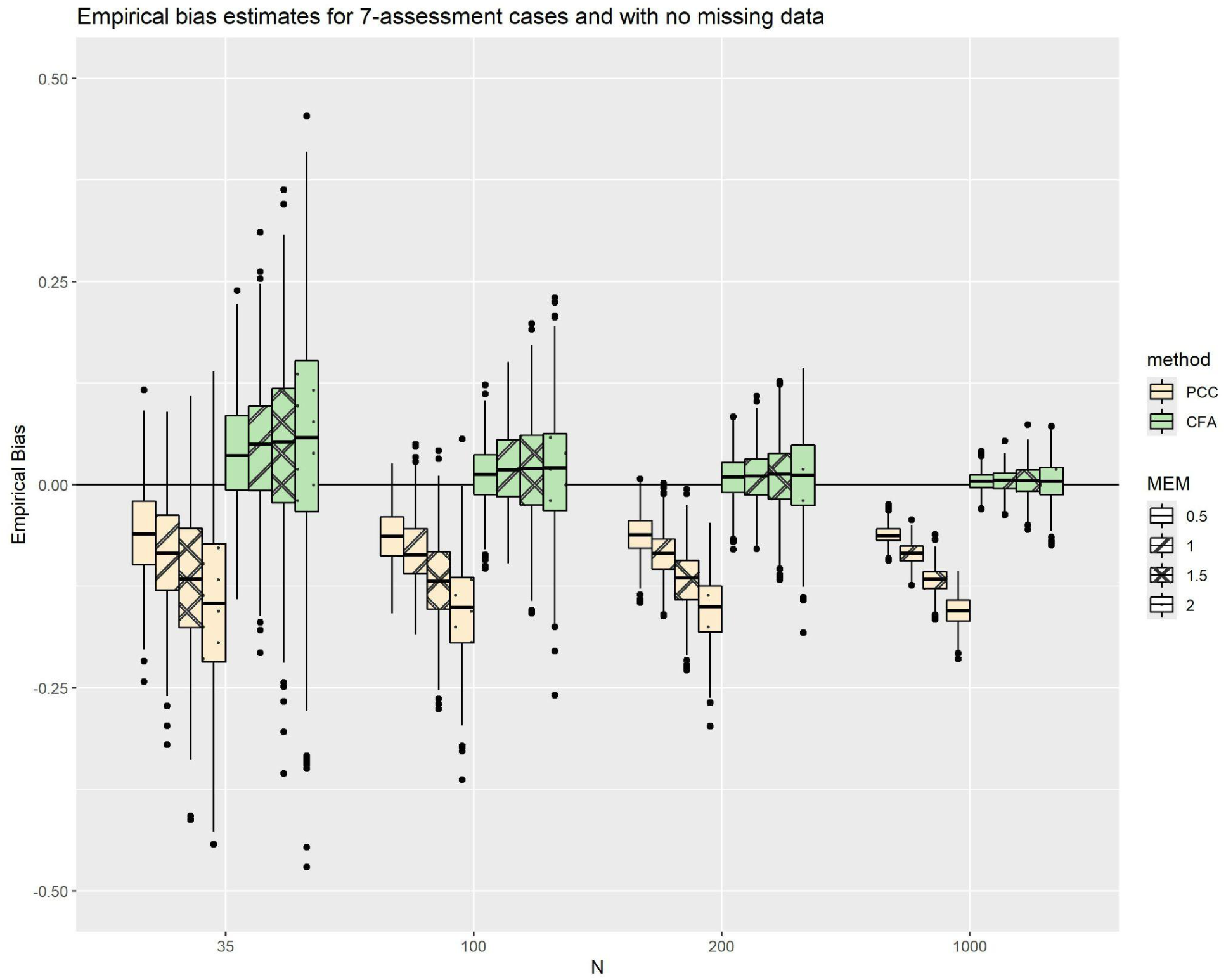
Empirical bias estimates for Pearson correlations and CFA factor correlations, as a function of increasing MEM, with no missing data and a fixed maximum number of repeated assessments

#### EmpSE

Table 3 shows that, overall, PCC had a lower mean empSE than CFA. In fact, a stronger condition is true - for every combination of simulation parameters investigated in the study, the empSE of CFA was greater than the corresponding empSE of PCC. This is also illustrated by Figure 5, which shows the empSEs of PCC plotted against the empSEs of the CFA factor correlation, colored by sample size.

**Figure 5.**
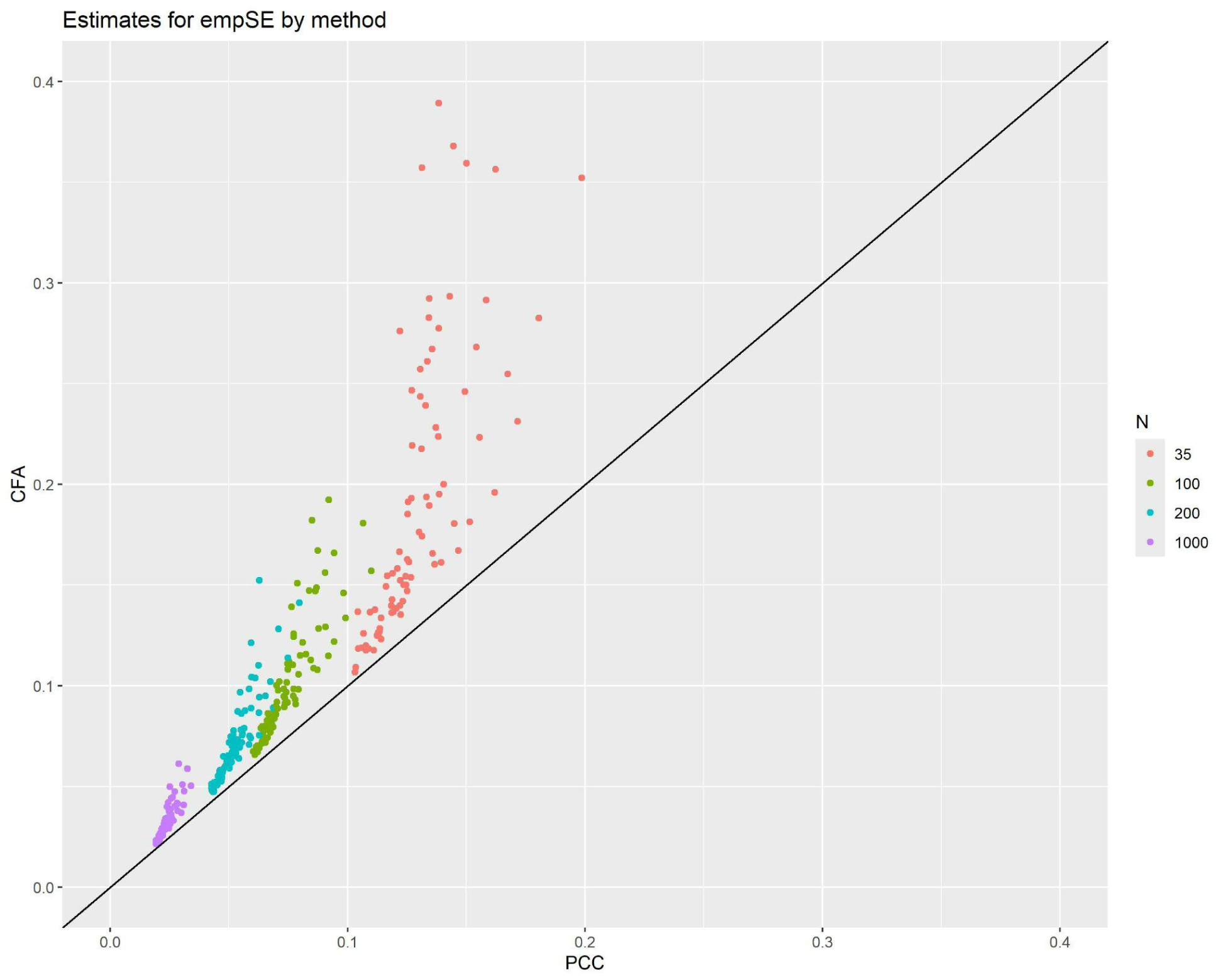
Plot of the empSEs for Pearson correlation against the empSEs for CFA factor correlation with line of identity. Colors represent the different sample sizes.

Continuing with Table 3: when considering the impact of any single simulation parameter on the mean empSE, decreasing the MEM or the missing rate caused the mean empSE to decrease for both PCC and CFA. For both methods, the empSE was lower with no missing data compared to that for MNAR data. The empSE for MNAR data was lower than that for MCAR data. When sample size increased, the mean empSE of both methods decreased. An increase in the number of assessments corresponded with a decrease in mean empSE for both methods. Such trends persist for more granular simulation scenarios, such as that shown in Table 6 (i.e., no missing data, seven repeated assessments).

**Table 6.** Empirical SE for each condition shown in Figure 5a.

| Sample size | MEM |  |  |  | Method |
| --- | --- | --- | --- | --- | --- |
| | $\frac{1}{2}\lambda_{\text{effect}}$ | $\lambda_{\text{effect}}$ | $\frac{3}{2}\lambda_{\text{effect}}$ | $2\lambda_{\text{effect}}$ | |
| 35 | 0.105 | 0.108 | 0.119 | 0.126 | Pearson |
|  | 0.118 | 0.118 | 0.136 | 0.191 | CFA |
| 100 | 0.061 | 0.066 | 0.066 | 0.070 | Pearson |
|  | 0.066 | 0.074 | 0.080 | 0.090 | CFA |
| 200 | 0.044 | 0.044 | 0.048 | 0.051 | Pearson |
|  | 0.047 | 0.049 | 0.058 | 0.067 | CFA |
| 1000 | 0.020 | 0.020 | 0.021 | 0.022 | Pearson |
|  | 0.022 | 0.023 | 0.026 | 0.029 | CFA |

Figure 7a depicts the empSEs for PCC and CFA factor correlations, as a function of increasing sample size with a fixed missing data rate of 0.40; Figure 7b depicts the corresponding empirical biases. When holding the missing data rate constant and allowing sample size to vary (Figure 7a), there was a pattern of increasing empSE with increasing MEM, which was mitigated by increasing sample size For example, the difference between the median empSE at the highest and lowest MEM levels in the PCC condition was 0.025 for n=35, whereas the difference between the highest and lowest MEM values was only 0.007 for n=1000 (i.e., a 3.6-fold difference). Furthermore, the empSE of CFA was more sensitive to increasing MEM at all sample sizes. The difference between the median empSE at the highest and lowest MEM levels was 0.202 and 0.018, for the n=35 and n=1000 cases respectively (i.e., an 11.2-fold difference). While the empSE of PCC also grew as MEM increased, its impact was less severe across all sample sizes. Empirical biases (Figure 7b) in these scenarios did not behave differently than the overall trend (Table 3).

**Figure 7a.**
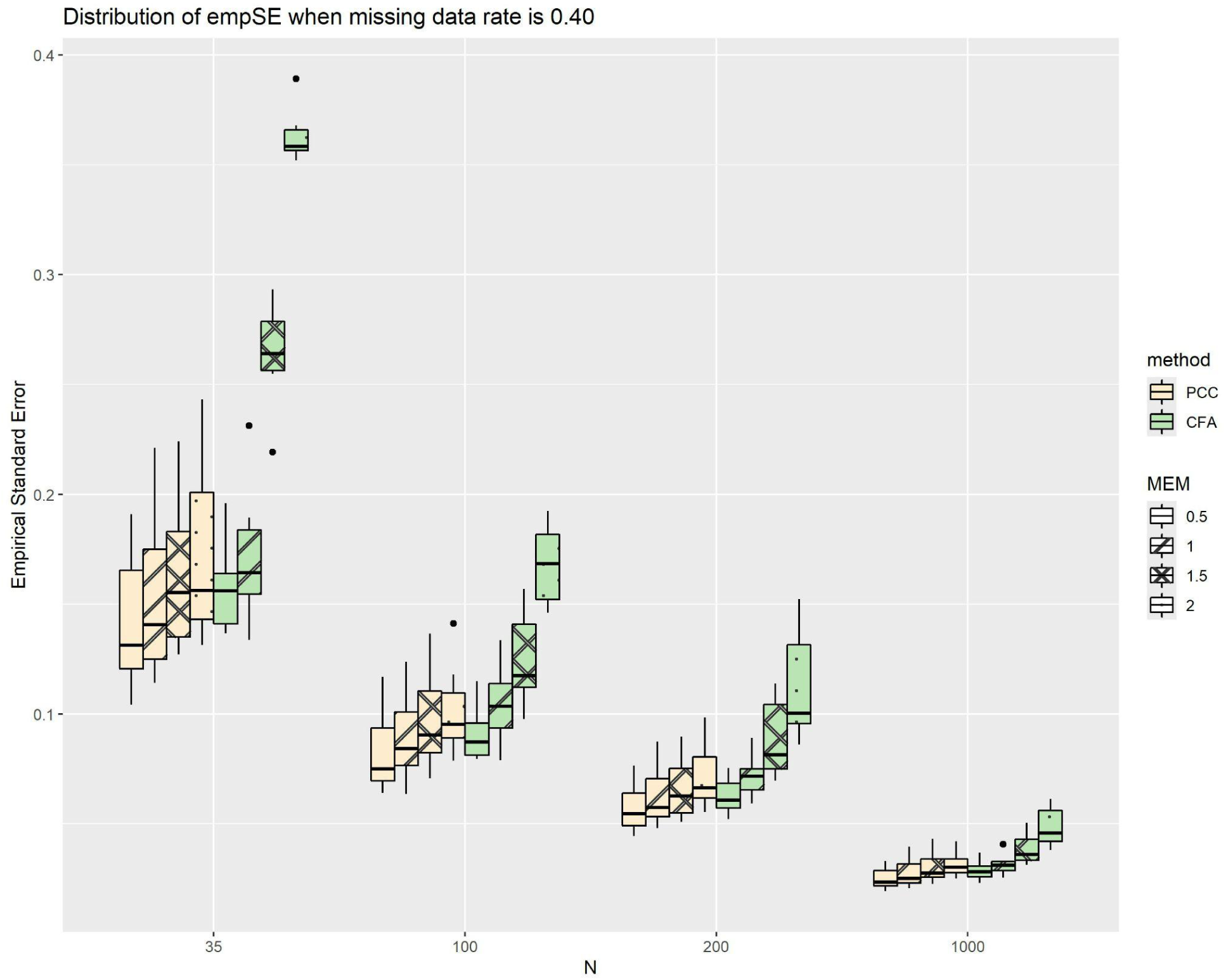
EmpSEs for both methods, as a function of increasing sample size with a fixed missing data rate of 0.40.

**Figure 7b.**
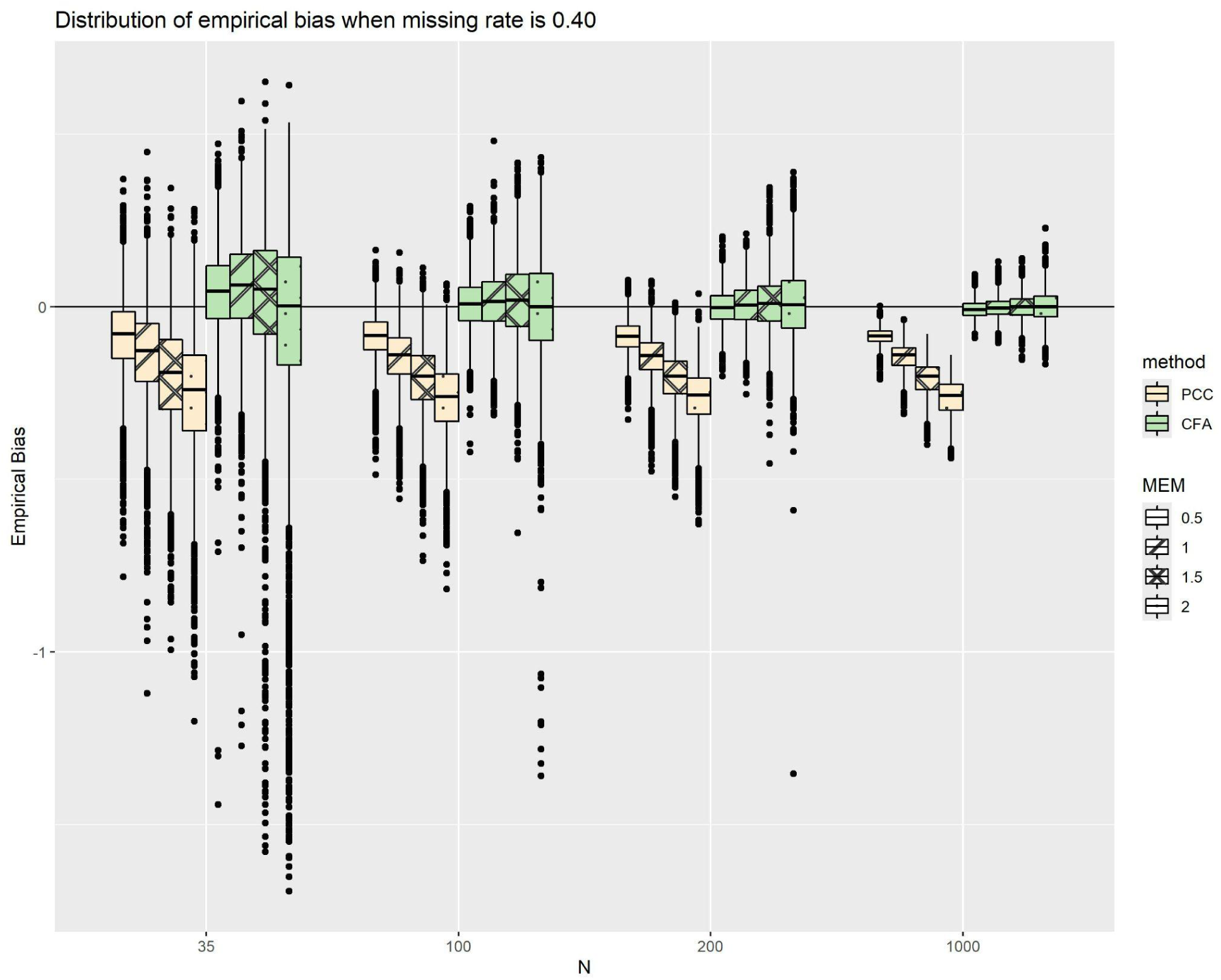
Empirical bias for both methods, as a function of increasing sample size with a fixed missing data rate of 0.40.

When holding sample size constant (N=35) and allowing MEM to vary, there was a trend for the empSE (Figure 8a) to perform notably worse in the 3-assessment cases than larger number of repeated assessments (RA = 5 and 7); this trend was mitigated by reducing the MEM, except that for largest simulated MEM (2λ_effect_) empSE for all RA were equally poor. While following a similar trend, PCC has notably smaller empSE across these scenarios as compared to CFA correlations. The same pattern holds in larger sample sizes (see supplementary materials 2, Figure 2a for N=100). Empirical biases (Figure 8b and supplementary materials 2, Figure 2b) did not differ notably from the overall trend (Table 3).

**Figure 8a.**
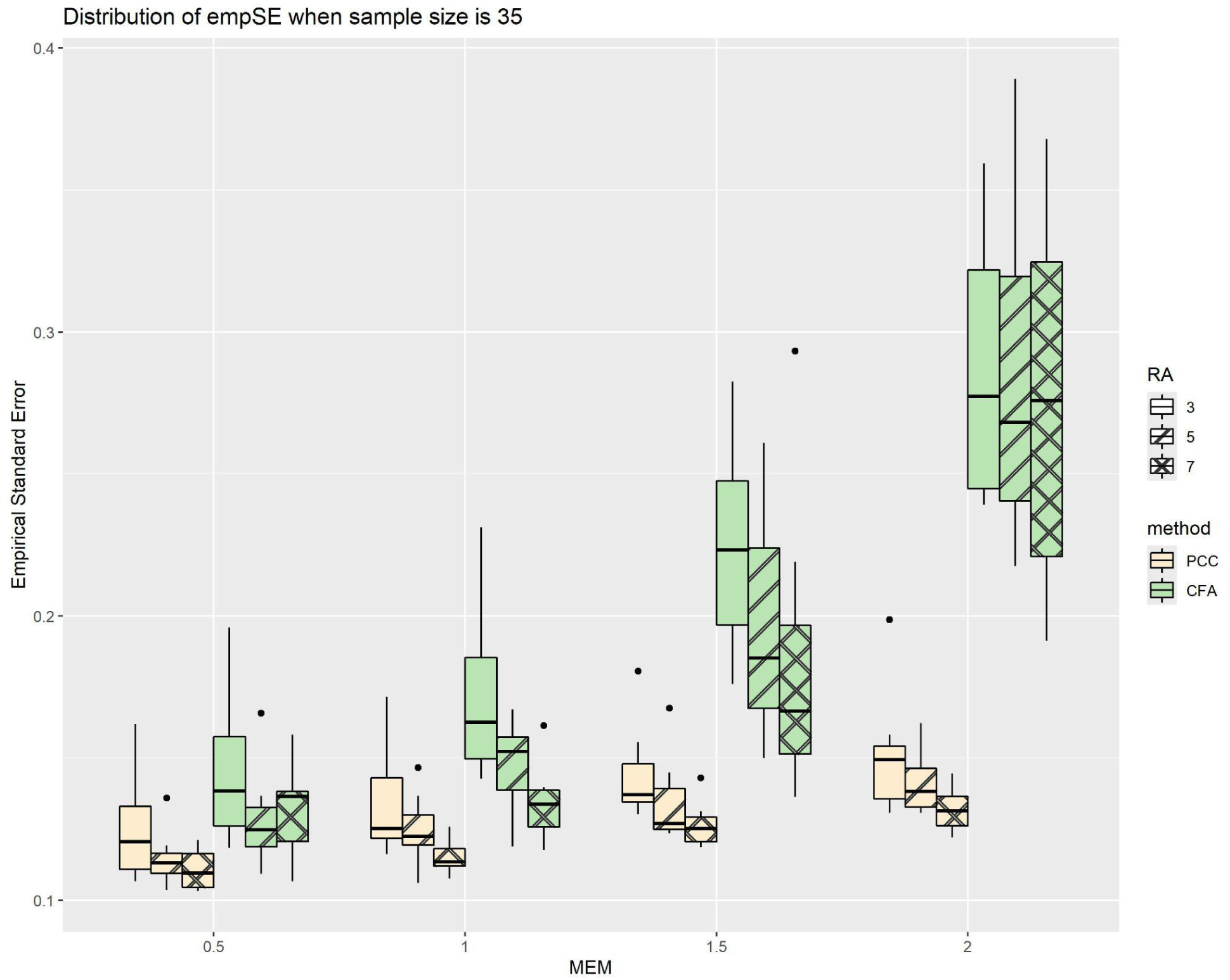
EmpSEs for both methods, as functions of MEM with a fixed sample size of 35.

**Figure 8b.**
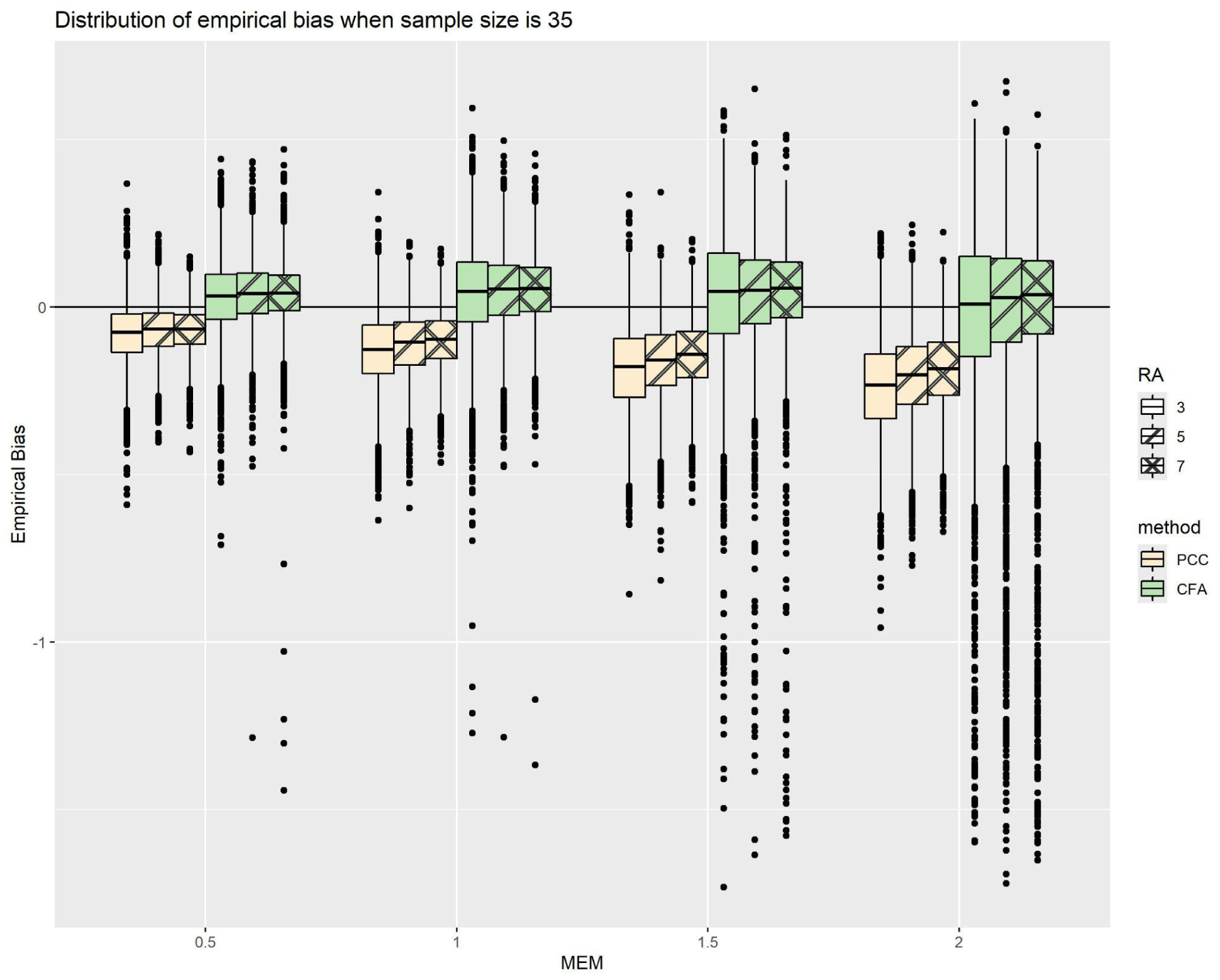
Empirical biases for both methods, as functions of MEM with a fixed sample size of 35.

#### CFA model fit statistics: rates of acceptable fit and failure rates

Table 9 depicts a summary of the rates of acceptable fit for each CFA model fit statistic and the model failure rate under the influence of different simulation conditions, as well as an overall summary across all conditions.

**Table 9.** Mean rates of acceptable fit for each model fit statistic, grouped in various ways. *acceptable fit is defined as those simulations with models with a CFI or TLI value of at least 0.9, and a RMSEA or SRMR value of less than 0.08; a failed model is one that resulted in an error output from the simulation code or returned a factor correlation not in the interval [-1,1].

|  | Mean (SD) rates of acceptable fit* |  |  |  | Mean (SD)<br>of Failure<br>rate<sup>#</sup> |
| --- | --- | --- | --- | --- | --- |
|  | CFI | TLI | RMSEA | SRMR |  |
| **Overall** | 84.8%<br>(28.1%) | 82.2%<br>(30.6%) | 45.4%<br>(44.6%) | 58.0%<br>(45.3%) | 2.67%<br>(5.74%) |
| By Sample Size |  |  |  |  |  |
| 35 | 59.1%<br>(36.8%) | 54.5%<br>(38.0%) | 20.0%<br>(31.8%) | <0.01%<br>(0.10%) | 10.1%<br>(7.48%) |
| 100 | 84.2%<br>(25.7%) | 80.3%<br>(29.3%) | 31.4%<br>(40.3%) | 34.7%<br>(30.6%) | 0.51%<br>(1.56%) |
| 200 | 96.0%<br>(12.0%) | 94.2% (15%) | 45.3%<br>(44.9%) | 97.2% (7.6%) | 0.06%<br>(0.30%) |
| 1000 | 100% (0%) | 100% (0%) | 84.7%<br>(30.8%) | 100% (0%) | 0% (0%) |
| By MEM |  |  |  |  |  |
| $\frac{1}{2}\lambda_{\text{effect}}$ | 99.6%<br>(1.30%) | 99.4%<br>(2.00%) | 89.9%<br>(18.5%) | 67.1%<br>(42.8%) | 1.37%<br>(2.47%) |
| $\lambda_{\text{effect}}$ | 96% (10.0%) | 94.6%<br>(12.7%) | 52.3%<br>(41.5%) | 60.3%<br>(44.4%) | 1.52% (2.8%) |
| $\frac{3}{2}\lambda_{\text{effect}}$ | 81.8%<br>(27.9%) | 78.2%<br>(30.8%) | 26.7%<br>(40.2%) | 54.3%<br>(46.1%) | 2.72%<br>(5.29%) |
| $2\lambda_{\text{effect}}$ | 61.9%<br>(37.7%) | 56.7%<br>(39.1%) | 12.6%<br>(28.9%) | 50.2%<br>(46.9%) | 5.07%<br>(9.07%) |
| By number of sDHT assessments |  |  |  |  |  |
| 1 | N/A | N/A | N/A | N/A | N/A |
| 3 | 88.9%<br>(21.5%) | 86.1%<br>(24.6%) | 55.3%<br>(43.3%) | 60.2%<br>(44.3%) | 3.83%<br>(7.86%) |
| 5 | 84.3%<br>(28.7%) | 81.6%<br>(31.4%) | 44.3%<br>(44.6%) | 57.5%<br>(45.7%) | 2.17%<br>(4.47%) |

|  | **Mean (SD) rates of acceptable fit\*** |  |  |  | *Mean (SD)*<br>*of Failure*<br>*rate<sup>#</sup>* |
| --- | --- | --- | --- | --- | --- |
|  | *CFI* | *TLI* | *RMSEA* | *SRMR* |  |
| 7 | 81.3%<br>(32.7%) | 79% (34.7%) | 36.5%<br>(44.2%) | 56.2%<br>(46.2%) | 2.01%<br>(3.97%) |
| By missing data mechanism |  |  |  |  |  |
| None | 91.4% (21.1%) | 89.5%<br>(23.8%) | 56.0%<br>(45.3%) | 65.0%<br>(43.6%) | 1.77%<br>(3.60%) |
| MCAR | 84.7%<br>(27.7%) | 82.0%<br>(30.2%) | 43.9%<br>(44.3%) | 56.4%<br>(45.6%) | 2.93%<br>(6.29%) |
| MNA<br>R | 82.8%<br>(30.3%) | 80.1%<br>(32.7%) | 43.3%<br>(44.4%) | 57.1%<br>(45.7%) | 2.71%<br>(5.76%) |
| By missing data rate |  |  |  |  |  |
| 0 | 91.4% (21.1%) | 89.5%<br>(23.8%) | 56.0%<br>(45.3%) | 65.0%<br>(43.6%) | 1.77%<br>(3.60%) |
| 0.10 | 89.1%<br>(24.0%) | 87.1%<br>(26.5%) | 51.7%<br>(45.3%) | 62.3%<br>(44.3%) | 2.08%<br>(4.22%) |
| 0.25 | 84.7%<br>(28.0%) | 82.0%<br>(30.4%) | 43.9%<br>(44.4%) | 57.3%<br>(45.7%) | 2.47%<br>(5.21%) |
| 0.40 | 77.3%<br>(33.3%) | 74.0%<br>(35.6%) | 35.1%<br>(41.9%) | 50.9%<br>(46.3%) | 3.91%<br>(7.92%) |

Across all conditions combined, fit statistics CFI and TLI were generally within acceptable bounds (84.8% and 82.2% respectively), and SRMR were generally acceptable but at a lesser level (58.0%). RMSEA were the least consistently acceptable, with only 45.4% meeting the criterion.

When considering the impact of varying any single simulation condition, increasing the parameters that control information quality (MEM and missing data rate) reduced the mean rate of acceptable fit across all fit statistics. MEM had a particularly large impact on RMSEA, decreasing the mean rate of acceptable fit by 67.3% as MEM increased from the minimum to the maximum value. The equivalent decrease in mean acceptable fit for SRMR was 16.9%, 37.7% for CFI and 42.7% for TLI. Varying the missing data mechanism had a much smaller impact on the mean acceptable rate for any of the fit statistics.

Increasing the sample size increased the mean acceptable fit rate for each statistic, strongly so in the case of RMSEA and SRMR. When sample size was 1000, each of CFI, TFI and SRMR always indicated an acceptable model fit, whereas RMSEA had an acceptable fit rate 84.7% of the time. When sample size was 35, almost none of the models were an acceptable fit according to SRMR, less than third were an acceptable fit according to RMSEA, and fewer than two-thirds were an acceptable fit according to CFI and TLI. Almost all of the unacceptable models according to SRMR occurred with a sample size of 100 or fewer.

Increasing the number of repeated assessments decreased the mean rate of acceptable fit for each fit statistic. The decrease in rate of acceptability was slight for each of CFI, TLI and SRMR, and of a slightly greater magnitude for RMSEA.

Figure 10 shows the rate of acceptable fit according to RMSEA and SRMR, restricted to N=100 and grouped by MEM. Figure 10 shows that the model fit according to RMSEA when N=100 was generally acceptable when MEM was at the minimum level of 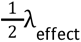; the mean acceptable fit rate was 91.2% compared to the 31.4% when considering all MEM conditions together for a sample size of 100. In contrast, almost no models had an acceptable fit according to RMSEA when MEM was at the maximum value - the mean acceptable rate was less than 1% in these cases as opposed to the overall 31.4% rate. SRMR acceptability rate exhibited a similar but weaker trend to RMSEA, with a mean acceptable rate of 68.4% at minimum MEM compared to 9.0% at maximum MEM (with an overall mean acceptable fit rate for SRMR when N=100 of 34.7%).

**Figure 10.**
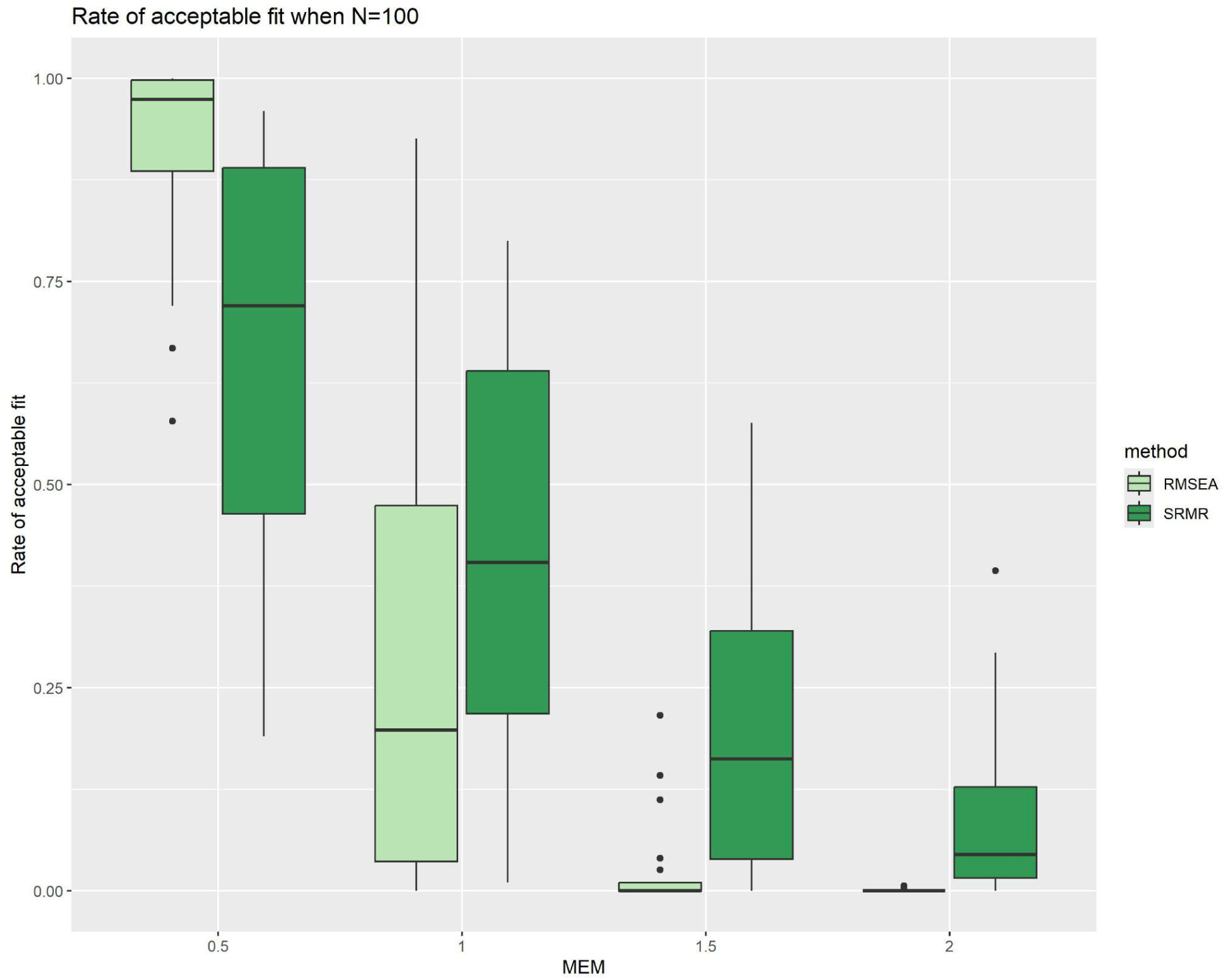
Rate of RMSEA and SRMR acceptable fit, grouped by MEM, when restricted to N=100

Similar but weaker patterns were seen for RMSEA and SRMR acceptable fit when grouping the N=100 cases by both the number of repeated assessments and by missing data rate: as the grouping parameter increased, the rate of acceptable fit decreased.

In approximately 9% of the total individual simulation repetitions, the RMSEA value was exactly zero. In all of these repetitions, the CFI was exactly 1.0 and the TLI was greater than 1.0. These repetitions were not limited to any particular combination of simulation conditions, although they were more likely to occur in conditions with lower levels of information (i.e., small number of assessments and small sample size) and better levels of signal to noise (i.e., small MEM and low missing data rate).

Almost all of the failures of the CFA model to converge occurred when sample size was 35, and sample size had the largest impact on failure rate when compared with other simulation parameters in this study. Increasing the MEM and missing data rate resulted in slight increases in failure rate. When including repeated target measure assessments, increasing the number of assessments led to a reduction in failure rate.

When attempting to use a single target measure assessment in the CFA model, the model was consistently found to be non-identifiable (i.e., multiple model specifications fit the data equally well). While factor correlations were computed in these cases, they have not been reported as they are unlikely to be meaningful.

### Secondary aim of the study

Table 11 depicts a summary of the mean change in information added *(ΔR*) and mean empSE for each regression method (SLR, MLR with “daily means”, MLR with “daily distinct”, and MLR with “weekly COAs”) under the influence of the different simulation conditions, as well as an overall summary across all conditions.

**Table 11.** Mean (SD) of ΔR2 and empirical SE for each regression model grouped in various ways.

| | Mean (SD) $\Delta R^2$ | | | Mean (SD) Empirical SE | | | |
| --- | --- | --- | --- | --- | --- | --- | --- |
|  | MLR -<br>“daily<br>means”<br>model | MLR -<br>“daily<br>distinct” | MLR -<br>“weekly<br>COAs”<br>model | SLR | MLR -<br>“daily<br>means”<br>model | MLR -<br>“daily<br>distinct”<br>model | MLR -<br>“weekly<br>COAs”<br>model |
| **Overall** | 0.198<br>(0.104) | 0.206<br>(0.123) | 0.031<br>(0.050) | 0.063<br>(0.036) | 0.068<br>(0.043) | 0.079<br>(0.064) | 0.069<br>(0.041) |
| By Sample Size |  |  |  |  |  |  |  |
| 35 | 0.180<br>(0.146) | 0.187<br>(0.190) | 0.014<br>(0.079) | 0.116<br>(0.015) | 0.129<br>(0.030) | 0.166<br>(0.068) | 0.130<br>(0.020) |
| 100 | 0.199<br>(0.098) | 0.207<br>(0.104) | 0.033<br>(0.043) | 0.068<br>(0.009) | 0.071<br>(0.013) | 0.076<br>(0.017) | 0.072<br>(0.010) |
| 200 | 0.204<br>(0.084) | 0.212<br>(0.086) | 0.038<br>(0.031) | 0.048<br>(0.007) | 0.049<br>(0.009) | 0.051<br>(0.011) | 0.050<br>(0.007) |
| 1000 | 0.208<br>(0.070) | 0.216<br>(0.072) | 0.042<br>(0.019) | 0.022<br>(0.003) | 0.022<br>(0.004) | 0.022<br>(0.004) | 0.022<br>(0.003) |
| By MEM |  |  |  |  |  |  |  |
| $\frac{1}{2}\lambda_{\text{effect}}$ | 0.280<br>(0.081) | 0.290<br>(0.092) | 0.048<br>(0.050) | 0.068<br>(0.038) | 0.059<br>(0.038) | 0.065<br>(0.05) | 0.071<br>(0.043) |
| $\lambda_{\text{effect}}$ | 0.221<br>(0.087) | 0.229<br>(0.104) | 0.036<br>(0.049) | 0.066<br>(0.037) | 0.067<br>(0.043) | 0.077<br>(0.060) | 0.070<br>(0.042) |
|  | MLR -<br>“daily<br>means”<br>model | MLR -<br>“daily<br>distinct” | MLR -<br>“weekly<br>COAs”<br>model | SLR | MLR -<br>“daily<br>means””<br>model | MLR -<br>“daily<br>distinct”<br>model | MLR -<br>“weekly<br>COAs”<br>model |
| $\frac{3}{2}\lambda_{\text{effect}}$ | 0.167<br>(0.089) | 0.173<br>(0.113) | 0.025<br>(0.048) | 0.062<br>(0.035) | 0.072<br>(0.045) | 0.085<br>(0.069) | 0.068<br>(0.041) |
| $2\lambda_{\text{effect}}$ | 0.125<br>(0.087) | 0.130<br>(0.117) | 0.016<br>(0.045) | 0.058<br>(0.033) | 0.072<br>(0.045) | 0.088<br>(0.075) | 0.065<br>(0.040) |
| By number of sDHT assessments |  |  |  |  |  |  |  |
| 1 | 0.154<br>(0.117) | 0.156<br>(0.152) | 0.014<br>(0.053) | 0.063<br>(0.038) | 0.079<br>(0.052) | 0.100<br>(0.090) | 0.072<br>(0.047) |
| 3 | 0.186<br>(0.098) | 0.209<br>(0.118) | 0.031<br>(0.049) | 0.064<br>(0.037) | 0.069<br>(0.043) | 0.079<br>(0.060) | 0.070<br>(0.042) |
| 5 | 0.219<br>(0.092) | 0.225<br>(0.102) | 0.039<br>(0.047) | 0.064<br>(0.035) | 0.063<br>(0.038) | 0.070<br>(0.048) | 0.067<br>(0.039) |
| 7 | 0.233<br>(0.088) | 0.233<br>(0.095) | 0.042<br>(0.045) | 0.062<br>(0.034) | 0.059<br>(0.035) | 0.065<br>(0.043) | 0.065<br>(0.037) |
| By missing data mechanism |  |  |  |  |  |  |  |
| None | 0.215<br>(0.096) | 0.223<br>(0.104) | 0.037<br>(0.045) | 0.058<br>(0.032) | 0.058<br>(0.035) | 0.064<br>(0.043) | 0.062<br>(0.036) |
| MCAR | 0.197<br>(0.108) | 0.205<br>(0.135) | 0.030<br>(0.054) | 0.068<br>(0.038) | 0.074<br>(0.048) | 0.089<br>(0.078) | 0.074<br>(0.045) |
| MNAR | 0.193<br>(0.102) | 0.200<br>(0.114) | 0.031<br>(0.046) | 0.06<br>(0.034) | 0.064<br>(0.039) | 0.073<br>(0.053) | 0.065<br>(0.038) |
| By missing data rate |  |  |  |  |  |  |  |
| 0 | 0.215<br>(0.096) | 0.223<br>(0.104) | 0.037<br>(0.045) | 0.058<br>(0.032) | 0.058<br>(0.035) | 0.064<br>(0.043) | 0.062<br>(0.036) |
| 0.10 | 0.208<br>(0.099) | 0.216<br>(0.108) | 0.035<br>(0.046) | 0.06<br>(0.034) | 0.062<br>(0.037) | 0.069<br>(0.047) | 0.064<br>(0.037) |
| 0.25 | 0.196 | 0.204 | 0.031 | 0.064 | 0.068 | 0.078 | 0.069 |
|  | MLR -<br>"daily<br>means"<br>model | MLR -<br>"daily<br>distinct" | MLR -<br>"weekly<br>COAs"<br>model | SLR | MLR -<br>"daily<br>means"<br>model | MLR -<br>"daily<br>distinct"<br>model | MLR -<br>"weekly<br>COAs"<br>model |
|  | (0.103) | (0.118) | (0.049) | (0.036) | (0.042) | (0.058) | (0.041) |
| 0.40 | 0.182<br>(0.111) | 0.188<br>(0.145) | 0.026<br>(0.055) | 0.068<br>(0.039) | 0.078<br>(0.051) | 0.096<br>(0.088) | 0.076<br>(0.048) |

#### Change in information added, *ΔR^2^*

Both MLR methods that included the daily PRO data performed well, recovering more information than the SLR model in the vast majority of cases (96.9% for the daily PRO means model, and 95.9% for the individual daily PRO days model). The MLR method including only the weekly COAs performed less well but was still generally better than the SLR model, adding more information than the SLR method in 77.8% of cases.

When considering the mean *ΔR*^2^, the MLR methods performed similarly well to each other, with a slight increase in mean *ΔR*^2^ for the “daily distinct” model as compared to the “daily means” model (Mean *ΔR*^2^ for “daily means” model = 0.206, mean *ΔR*^2^ for “daily distinct” model = 0.198) The “weekly COAs” model performed much worse, with a mean *ΔR*^2^ of 0.031. This pattern was repeated when considering the results grouped by any single simulation parameter.

When grouping the *ΔR*^2^ results by any single simulation parameter, the largest impacts on *ΔR*^2^ were observed to be caused by MEM and the number of repeated assessments. In general, increasing the sample size and number of assessments led to an increase in *ΔR*^2^ for all models. Moreover, reducing the MEM and missing data rate also increased *ΔR*^2^ for all models. Sample size had a particularly notable impact on the variability of *ΔR*^2^ under the daily distinct model, with a SD of 0.190 at N=35 compared to 0.072 at N=1000. Values of *ΔR*^2^ from simulations with a sample size of 35 exhibited the greatest variability of all models across all conditions.

When investigating simulations with negative *ΔR*^2^ values (3.1% of all values for the means model, 4.1% of all values for the daily distinct model, and 22.2% of all values for the weekly COAs model), sample size had the greatest impact out of any single simulation parameter. When sample size was 35, over half of the *ΔR*^2^ values for the weekly COAs model were negative, decreasing to around a quarter of simulations at N=100, then around 10% at N=200, and almost no simulation at N=1000 produced a negative *ΔR*^2^ for this model. This pattern was repeated for the other two models; between 10% and 15% of simulations at N=35 returned a negative *ΔR*^2^ for the means and daily distinct models respectively, which reduced to almost zero for larger sample sizes.

There were a limited number of simulations where the MLR *R* values were negative; the majority of these negative values occurred when sample size was 35.

#### EmpSE

Overall, the empSE was similar between all MLR methods and the SLR method. The SLR method had a slightly smaller mean empSE, with the “daily means” model and “weekly COA” models having slightly larger but broadly similar means, and the “daily distinct” model having the largest mean empSE among all four methods.

When considering any single simulation parameter, varying the sample size resulted in the strongest impact on empSE, with an increase in sample size tending to result in a decrease in empSE for each regression method.

This trend was less clear when varying the number of repeated assessments. For MLR methods there was a slight trend of decrease in mean empSE as the number of assessments increased, but for the SLR method, varying the number of assessments had broadly no impact.

Introducing missing data via MNAR tended to produce a slightly reduced empSE in each method as compared to MCAR, and increasing the missing data rate tended to slightly increase empSE for each method.

When allowing just the MEM to vary, there was a more complicated picture. For both models that included the daily PRO, empSE exhibited a slight tendency to increase as MEM increased. However, for methods only including weekly COAs (i.e., the SLR and the MLR “Weekly COAs model) this was reversed, and the empSE exhibited a slight tendency to decrease as MEM increased.

#### Regression model failure rates

There were a limited number of failures – that is to say, that NaN was returned as the 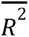 estimate - in the “daily distinct” MLR model, all of which occurred when sample size was 35 and missing data rate was at its maximum of 0.40 under the MCAR mechanism. Table 12 lists the specific simulation parameter combinations that led to failures, along with their failure rates.

**Table 12.** Conditions where the “day-by-day” model produced failures, along with the failure rate.

| **N** | **MEM** | **Number of**<br>**sDHT**<br>**assessments** | **Missing**<br>**data**<br>**mechanism** | **Missing**<br>**data rate** | **Failure rate** |
| --- | --- | --- | --- | --- | --- |
| 35 | 0.5 | 1 | MCAR | 0.4 | 1.85% |
| 35 | 1 | 1 | MCAR | 0.4 | 2.24% |
| 35 | 1.5 | 1 | MCAR | 0.4 | 1.69% |
| 35 | 2 | 1 | MCAR | 0.4 | 2.57% |
| 35 | 1.5 | 3 | MCAR | 0.4 | 0.18% |

There were a small number of 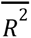 estimates produced from the daily distinct model which strongly underperformed the SLR *R*^2^; these estimates tended to arise in the same simulation conditions where model failures were reported.

The SLR model, the “daily means” MLR model, and the “weekly COAs model exhibited no failures during the simulation.

#### Monte Carlo standard errors of the empSE

All MCSE values across each of the simulation conditions were smaller than the acceptability criteria of 0.025, for each of the six statistical methods investigated in this work. Table 13 shows the mean MCSE of the empSE for each method, and it can be observed that these values are all comfortably below the acceptability threshold.

**Table 13.**
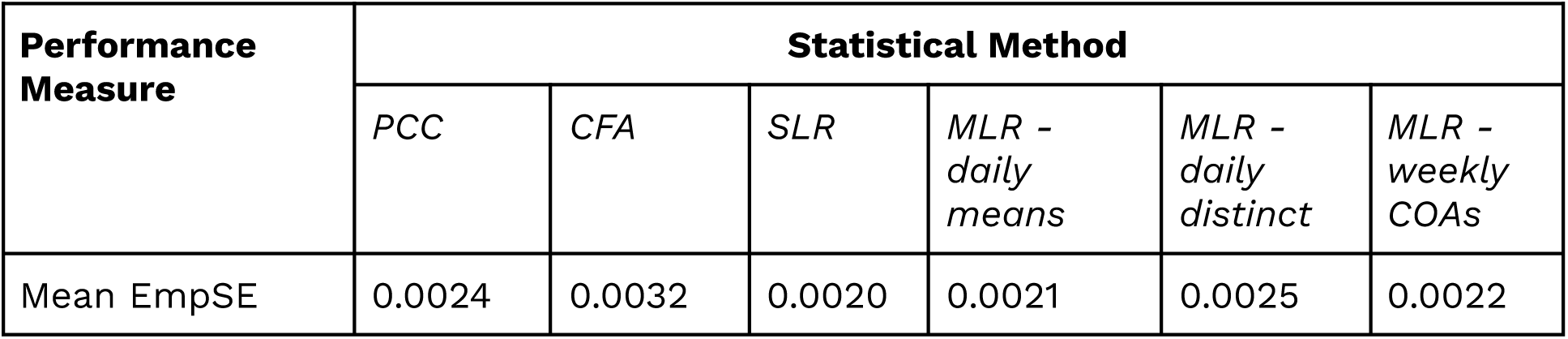
MCSEs of the mean empSEs for each statistical method investigated.

| Performance Measure | Statistical Method |  |  |  |  |  |
| --- | --- | --- | --- | --- | --- | --- |
|  | *PCC* | *CFA* | *SLR* | *MLR - daily means* | *MLR - daily distinct* | *MLR - weekly COAs* |
| Mean EmpSE | 0.0024 | 0.0032 | 0.0020 | 0.0021 | 0.0025 | 0.0022 |

## Discussion

### Summary of results and how they help in practice

In this simulation study, we conducted AV of a sDHT against a single reference measure that lacks direct correspondence. We did so by comparing Pearson correlations and CFA factor correlations. Variables and parameter choices were informed by real-world data and observation, as were some of the missing data mechanisms.

The Pearson correlation was more stable, easier to compute, and relatively robust with respect to violations of parametric assumptions [26], making it the recommended choice in small sample size studies (i.e., N=35). However, Pearson correlations tend to underestimate the true correlation, and such underestimation is magnified as the degree of measurement error increases.

CFA factor correlation has increased in popularity among researchers as a viable solution because its approach to extracting latent information can better mitigate the statistical impacts of measurement error on estimation. However, complex models such as CFA require a well-sized sample (N>=100) and intensive repeated assessments from the sDHT. The CFA model was shown to be unsuitable for a single assessment scenario due to model non-identifiability. Additionally, model fitting evaluation needs to be carefully investigated. In our simulated examples, commonly used fit statistics CFI and RMSEA tended to yield different qualitative conclusions.

Specifically, CFI indicated acceptable model fits for >80% of simulated cases while results from RMSEA were in agreement around half of the time (Table 9). The mixed results do not necessarily indicate model misspecification; reasons for the disagreements may include small sample size and violation of parametric assumptions [27]. We provided results from all these statistics to give readers a complete picture of CFA fitting in our simulated data. Further research is needed to investigate behaviors of CFA fit statistics in commonly occurring sDHT data distributions.

Strengths and weaknesses of Pearson correlation and CFA factor correlation complement each other. Researchers should consider presenting results from both methods when assessing convergent validity. Pearson correlation, a conservative and stable method, demonstrates the relationship between target and reference measures and may provide a lower bound of true correlation. On the other hand, CFA factor correlation provides an optimistic but intuitive estimate, serving as an upper bound. The true correlation likely lies between the results from these two methods.

Additionally, including multiple reference measures at a higher time resolution (e.g., values recorded daily instead of weekly) can bring benefits. Specifically, multiple reference measures can help increase the information available for mapping the sDHT to these reference measures, providing a more complete presentation of the sDHT’s AV. Using an MLR approach in this work with daily collected reference measures notably boosted the explainable variance from the sDHT, compared to when the reference measure was only collected once.

### Novelty of the work

We operationalized the AV of a sDHT in a physical activity scenario in terms of psychometric concepts (i.e., latent and observable variables). Our procedure provides investigators with a general-use simulation-based framework for assessing how well a sDHT relates to clinical outcomes. It can be applied to various domains such as speech-based measures of rate, or cognitive-linguistic measures.

Investigators can use our simulation study procedure to investigate various endpoints (primary, secondary, surrogate, etc.), such as freezing of gait episodes in Parkinson’s Disease, or depression severity using passively collected data from mobile devices [28]. Following this study, the simulation code, simulated data, full results, DGM, and guidance of how to adapt these materials to different digital measure scenarios, will be made available open-access through the website of the Digital Medicine Society [29] and a public data repository [30] for investigators to adapt as needed.

CFA factor correlation was evaluated as a way to handle data believed to be generated by latent traits, together with the Pearson correlation, in the context of AV of sDHTs. We believe our study is the first to take this approach. Our results provide researchers with crucial information for choosing appropriate methods to conduct AV.

### Limitations and future research

Future work may consider how to specify the DGM in a way that explicitly controls the true relationship between the sDHT and the reference measure(s), e.g. explicitly controlling the true correlation between the measures. This may allow a researcher to specify a single parameter independent of simulation repetitions that the statistical models can attempt to identify. They could therefore, for example, estimate the true bias of each model, instead of using empirical bias as the current work has.

When it comes to modeling missingness, the MCAR mechanism was chosen to recreate missing data patterns observed in the collection of physical activity data during clinical trials [31]. In the simulation, whether values were MCAR or MNAR did not substantially affect the performance of any of the methods. Further work could explore the impact of other missing data mechanisms on the performance of the statistical methods: in particular, mechanisms that more closely model observed real-world trends. These might include mechanisms applied at the epoch level as opposed to the summary level, and an inverted MNAR method (that is, to make smaller data points more likely to be deleted than larger ones) [32]. Additionally, missing data was simulated only for the target measure, and future work should explore how missing reference measure(s) data may impact the analysis results.

Lastly, statistical methods investigating errors-in-variables, such as Deming regression, may be explored. Errors-in-variables models would be particularly suitable for use in this scenario due to their accounting for error in the predictor variables.

Another method that warrants investigation is CFA when modeling multiple reference measures. A potentially suitable correlated two-factor model could be thus: the items from the weekly COAs, and the daily assessments from the daily PRO, are loaded onto separate factors. These factors are then loaded onto a combined “reference measure” factor, which is then correlated against a “target measure” factor with each day of sDHT data loaded as separate indicators. The two-factor CFA model used in this work might also be modified in future, to investigate if the model fit may be improved for small sample sizes and other conditions. For example, one might introduce covariances between consecutive days of target measure data - an approach which more closely matches the design of the DGM.

## Acknowledgements

The authors gratefully acknowledge the contributions of the following experts through participation in the statistical advisory committee and async review of the simulation protocol and results: Chakib Battoui, Jakob Bjørner, Yiorgos Christakis, Valentin Hamy, Andrew Potter, Bohdana Ratitch, David Reasner, Colleen Russell, Sachin Shah, Berend Terluin, Andrew Trigg, Kevin Weinfurt, Robert Wright. In addition, the authors gratefully acknowledge the contributions of DiMe members for their support: Samantha

## Data availability statement

Simulated data used in this study, along with the R code used to create and analyze the simulated data, will be made available through the website of the Digital Medicine Society (https://datacc.dimesociety.org/validating-novel-digital-clinical-measures), and through the Open Science Framework (https://osf.io/3shq8).

## Declaration of conflicting interests

ST is a contractual employee of the Digital Medicine Society.

LS has no conflicts to report.

JL has no conflicts to report (Activinsights Ltd. & University of Exeter).

CC reports employment and equity ownership in Verily Life Sciences (ORCID: 0000-0003-1175-3027).

RA has no conflicts to report.

PG is a contractual employee of the Digital Medicine Society and President of SeeingTheta.

LF has no conflicts to report.

EJD has no conflicts to report. (ORCID: 0000-0002-8376-1600)

