## Supplementary materials for "Methods for analytical validation of novel digital clinical measures: A simulation study"

#### S1 - Further details on the simulation parameters

##### Note on the missing data mechanisms

Missing data mechanisms were only applied to the target measure data. To simulate intermittent (i.e., non-monotonic) MNAR, data points were selected for deletion based on the magnitude of the data point for that day - the higher the value of that day's target measure value, the more probable that datapoint was selected for deletion. More specifically, the ratio of data points above the overall median weekly value that were deleted compared to data points at or below the median value that were deleted was 4:1.

To simulate MCAR, data points were selected for deletion in two ways. FIrstly, data points on the first day of the study were deleted completely at random, to model logistical issues such as late delivery of the device and late start of the participant. Additionally, to model monotonic missingness resulting from device failures and drop-out due to low adherence, data on consecutive days were deleted completely at random based on an exponential distribution. The generated value from the exponential distribution for a given individual indicated after which day in the trial their data should be deleted. The exponential parameter value varied based on the MDR being simulated ($\lambda_{exp} = \frac{1}{42}$ for MDR of 0.10, $\lambda_{exp} = \frac{1}{15}$ for MDR of 0.25, $\lambda_{exp} = \frac{1}{8}$ for MDR of 0.40).

##### Note on the implementation of fewer repeated assessments

When including fewer than seven target measure assessments in the simulation, consecutive days were removed starting from the beginning of the study; for example, when RA=3, only target assessment data from days 5, 6, and 7 were included, and only data from those days contributed to the true mean of the target measure used in calculating the PCC and regression models. Further, the CFA model loads only those days as indicators onto the “target measure” factor.

All seven days of the daily PRO data were included in the MLR models, regardless of the RA value.



Unpublished manuscript

### S2 - Further empSEs and empirical biases of correlation methods

Figure 1 depicts empSEs for Pearson correlations and CFA factor correlations as a function of increasing number of sDHT assessments. Figure 1a further groups the values by sample size, and Figure 1b further groups the values by MEM.

When grouped by number of repeated assessments and sample size (Figure 1a), average empSE decreased as the number of repeated assessments increased for all cases with varying sample sizes, and this trend was more pronounced at smaller sample sizes. When grouped by number of repeated assessments and MEM (Figure 1b), average empSE increased as the magnitude of MEM increased, and the trend was more pronounced at the highest MEM value than at the lower MEM values.

manuscript

**Fig 1a - Empirical SEs for both methods, grouped by number of repeated assessments and sample size**

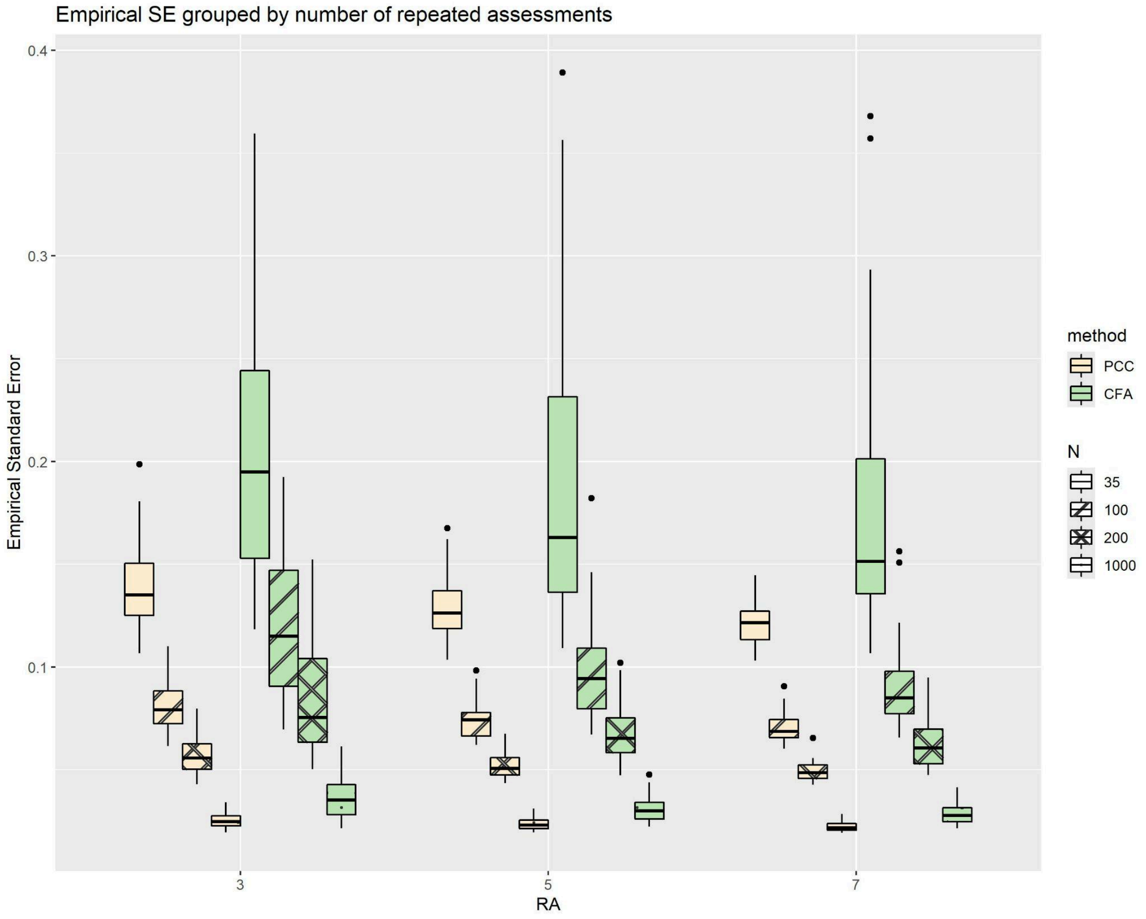

Fig 1b - Empirical SEs for both methods, grouped by number of repeated assessments and MEM

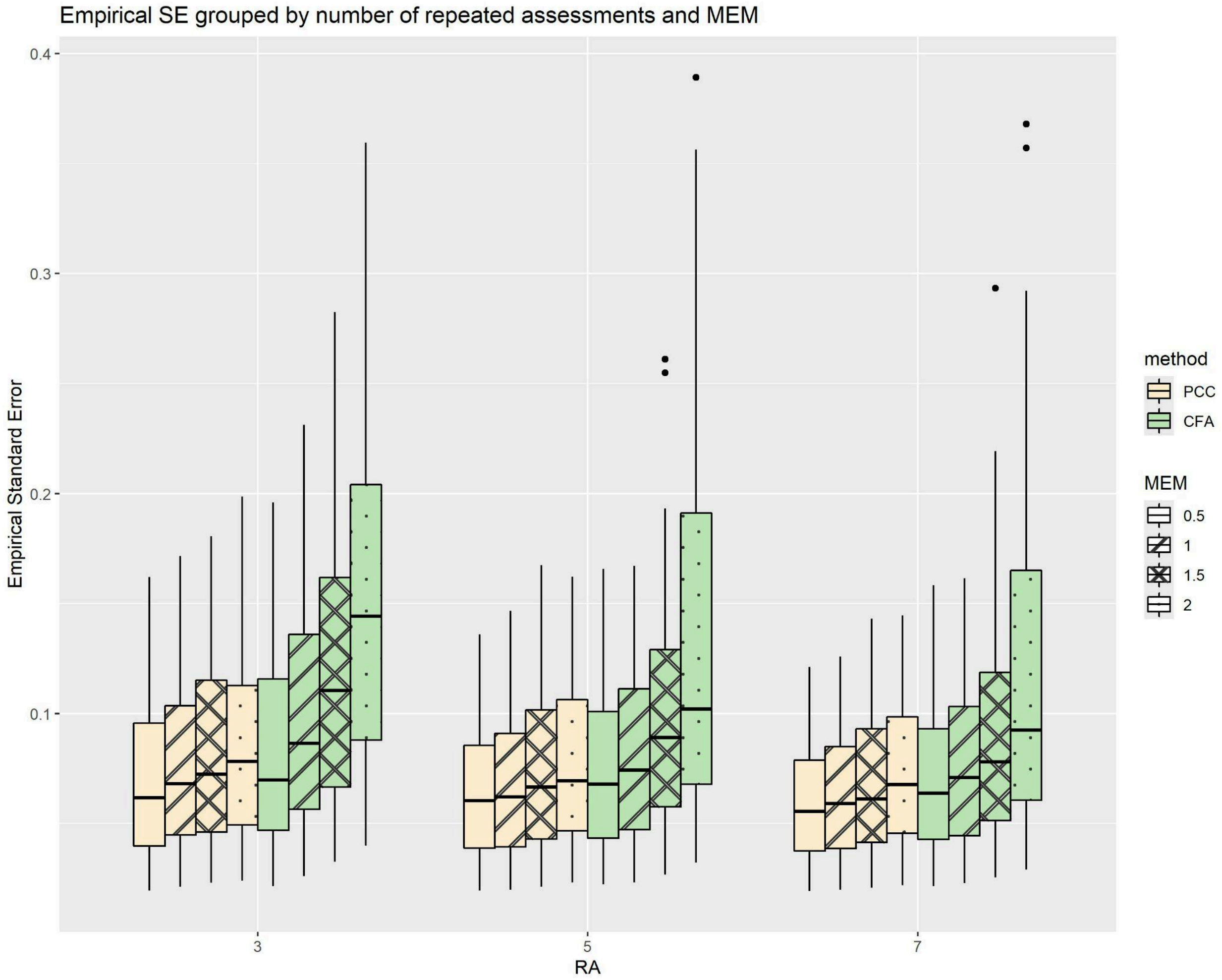


Unpublis

Figure 2a depicts the empSEs for PCC and CFA factor correlations, as a function of increasing MEM with a fixed sample size of 100; figure 2b depicts the corresponding empirical biases. Both figures 2a and 2b illustrate that the observed trends for N=35 cases (Main text, figures 8a and 8b) are not changed when allowing the sample size to increase.

**Fig 2a - EmpSEs for both methods, as functions of MEM with a fixed sample size of 100.**

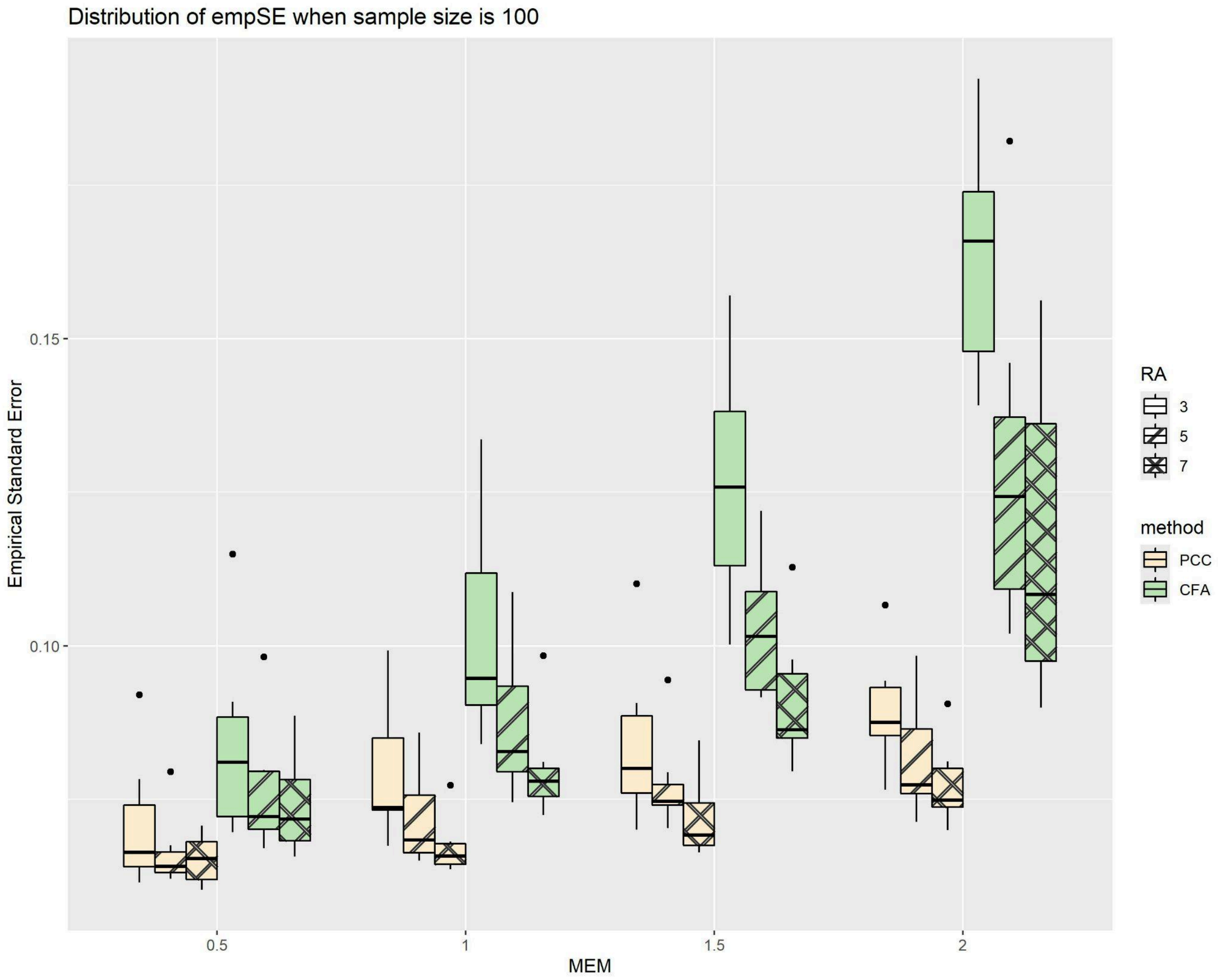

Fig 2b - Empirical biases for both methods, as functions of MEM with a fixed sample size of 100.

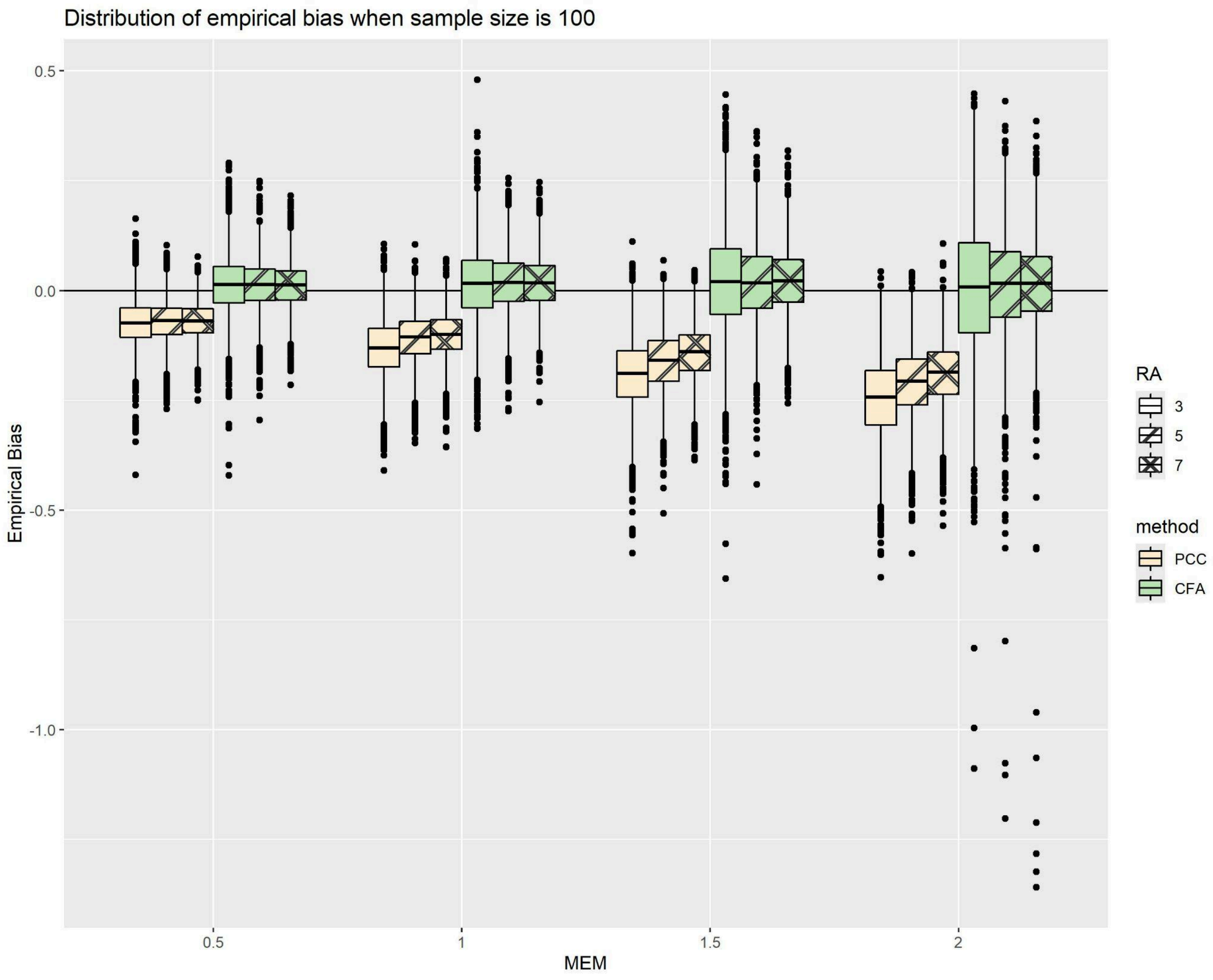


Unpubli
